# Predicting *Salmonella* Typhi incidence using prevalence metrics from sentinel studies of community-onset bloodstream infections

**DOI:** 10.64898/2026.02.13.26346225

**Authors:** Nienke N. Hagedoorn, Shruti Murthy, Christian S. Marchello, Jonathan Williman, Faisal Ahmmed, Jason R. Andrews, Buddha Basnyat, Alice S. Carter, Shrimati Datta, Irum Fatima Dehraj, Kate Doyle, Denise O. Garrett, Jobin Jacob, Hyonjin Jeon, Jacob John, Farhana Khanam, Jooah Lee, Xinxue Liu, Florian Marks, Shiva R. Naga, Kathleen Neuzil, Paul N. Newton, Priyanka D. Patel, Andrew J. Pollard, Firdausi Qadri, Farah Naz Qamar, Tamalee Roberts, Jessica C. Seidman, Mila Shakya, Suchita Shrestha, Birkneh T. Tadesse, Dipesh Tamrakar, Manivanh Vongsouvath, Merryn Voysey, Mohammad Tahir Yousafzai, John A. Crump

## Abstract

**Background:** Typhoid fever incidence estimates are central to policy decisions on vaccine introduction and investments in non-vaccine prevention and control but are often unavailable. We explored whether prevalence metrics from sentinel studies of community-onset bloodstream infections could accurately predict local *Salmonella* Typhi (*S.* Typhi) incidence.

**Methods:** Using a previous systematic review (January 2018-December 2024), we identified studies reporting both typhoid incidence and prevalence of community-onset bloodstream infections from sentinel sites. From authors, we requested data on blood culture isolates and analysed four metrics: (i) *S.* Typhi prevalence among probable pathogens, (ii) *S.* Typhi rank order, (iii) *S.* Typhi to *Escherichia coli* ratio, and (iv) *S.* Typhi to ‘stably endemic’ organisms ratio. Typhoid incidence was categorized as low (<10), medium (10-100) or high (>100) per 100,000 person-years. We used univariate ordinal regression to assess the association between each metric and typhoid incidence level. The model performance was evaluated by the c-statistic, sensitivity, and specificity.

**Findings:** Analysis of 29 study sites (20 Africa, 9 Asia) yielded 4,625 probable pathogens. The median (IQR) typhoid incidence was 140 (28-319) per 100,000 person-years. All metrics were associated with increased typhoid incidence level: for each 1% increase in *S.* Typhi prevalence OR 1.07 (95%CI 1.02-1.15); rank order OR 0.25 (95%CI 0.06-0.64); log *S.* Typhi to *E. coli* ratio OR 2.91 (95%CI 1.45-7.42); log *S.* Typhi to ‘stably endemic’ organisms ratio OR 3.69 (95%CI 1.69-11.3). A parsimonious model using *S.* Typhi prevalence alone achieved c-statistics of 0.87 (0.58-0.97), 0.76 (0.51-0.91), and 0.88 (0.69-0.96) for low, medium, and high incidence, respectively.

**Interpretation:** Sentinel prevalence metrics from bloodstream infections, particularly *S.* Typhi prevalence among probable pathogens, could be useful for inferring local typhoid fever incidence where direct data are unavailable.

**Funding:** Gates foundation

**Research in context:** *Evidence before this study:* Globally, annual deaths from typhoid fever were estimated at 71,954 (95% uncertainty interval 38,051 to 118,560) in 2023. Typhoid conjugate vaccines (TCV) are recommended for regions with high typhoid incidence. Implementation, however, can be challenging due to a lack of local incidence data. Generating community incidence estimates requires expensive and time-consuming large prospective or hybrid surveillance studies, or novel techniques such as serology or environmental surveillance. Our previous study proposed that metrics from sentinel healthcare facilities such as the prevalence of *Salmonella* Typhi (*S.* Typhi) among all bloodstream pathogens or its rank order relative to other pathogens could serve as proxy for community incidence. However, contemporaneous incidence and prevalence data from the same time and location were limited in our previous study. To explore typhoid incidence estimation strategies, we searched PubMed and MEDLINE on January 8, 2026 with search terms including keywords of “typhoid fever”, “incidence”, and “prediction” without restrictions to language or publication date. Previous studies estimated incidence based on complex country-level covariates and disease modelling that lack ease of applicability for policy decisions. Recognising the need for pragmatic tools, we explored whether prevalence metrics from sentinel studies of community-onset bloodstream infections could accurately predict local *S.* Typhi incidence.

*Added value of this study:* Our study was based on typhoid incidence studies that had available data for isolates of bloodstream infections. Of 29 sites across Africa and Asia with 4,625 probable pathogens, we found that all four sentinel metrics were significantly associated with typhoid incidence level. We demonstrated that a parsimonious model using *S.* Typhi prevalence alone achieved good discriminative performance in identifying high incidence settings.

*Implications of all the available evidence:* When typhoid incidence estimates are unavailable, prevalence metrics from sentinel studies of community-onset bloodstream infections could help policymakers infer typhoid incidence and optimise resource allocation in water, sanitation, and hygiene, and TCV introduction.

## Background

Invasive *Salmonella* disease occurs when *Salmonella* spp. invades normally sterile sites and is often associated with severe illness. Typhoid fever is caused by *Salmonella enterica* subspecies *enterica* serovar Typhi (*S.* Typhi). Worldwide, annual deaths from typhoid fever were estimated at 71,954 (95% uncertainty interval 38,051 to 118,560) in 2023.^1^ Prequalified typhoid conjugate vaccines (TCVs) are available and recommended for use by the World Health Organisation to prevent disease.^2,3^ Accurate estimates for typhoid incidence are central to policy decisions on vaccine introduction and, additionally, investments in non-vaccine prevention and control. However, generating estimates for local typhoid incidence usually require large, costly, and time-consuming prospective active or hybrid surveillance studies.

We previously proposed that metrics from sentinel studies that monitor community-onset bloodstream infections in healthcare facilities, henceforth called ‘prevalence studies,’ are considerably less resource-intensive and more widely available than population-based active or hybrid surveillance studies, and could aid in making inferences of local typhoid incidence. ^4^ These prevalence studies offer potential metrics, such as the prevalence of *S.* Typhi among bloodstream pathogen isolates, its rank order relative to other pathogens, or prevalence relative to endemic organisms such as *Escherichia coli (E. coli)*. In our previous study, concurrent data from the same location for typhoid incidence and the prevalence metrics of bloodstream infections were few, limiting our ability to make strong inferences.^4^

Recently, large scale surveillance studies and vaccine efficacy trials across Africa and Asia have obtained robust data necessary to validate sentinel prevalence metrics.^5–11^ We examined the relationship of *S.* Typhi incidence with metrics from sentinel community-onset bloodstream infections in order to propose a pragmatic tool for policy decisions for control of typhoid fever.

## Methods

### Study design and data sources

We performed a secondary analysis of data collected by global studies that reported typhoid incidence data and that additionally had data available for all individual bloodstream isolates. First, we identified eligible reports through an updated systematic review of global typhoid incidence by Murthy et al.,^12^ focusing on studies published between January 1, 2018, and December 15, 2024, and we consulted experts involved in typhoid fever research for additional reports meeting our criteria. To be included, studies were required to report typhoid incidence and had data available on the prevalence of community-onset bloodstream infection at sentinel sites. For identified studies where data on bloodstream isolates were missing from published manuscripts or linked databases, we requested primary blood culture data directly from the study authors. If the study was a typhoid vaccine trial, we requested data stratified by individual typhoid vaccine immunisation status, or by cluster in case of a cluster-randomised trial.

### Data abstraction and data requests

We abstracted from the included articles information on study design, the period of data collection, location of data collection, age range of included participants, blood culture criteria, and laboratory methods on blood culture processing and isolate identification. We abstracted the typhoid incidence estimate and 95% confidence interval (CI) or uncertainty range, and when reported, typhoid incidence estimates stratified by age. For vaccine trials, we abstracted incidence estimates for the control arm of the trial. From study authors, we requested all blood culture results including the genus, species, and for *Salmonella enterica* serotype of isolates, date of blood culture, age at blood culture, and hospitalisation status of participants.

### Definitions

We classified all isolates as either likely contaminants or probable pathogens. We defined likely contaminants as non-*anthracis Bacillus* species, coagulase-negative staphylococci, *Corynebacterium* species, and *Micrococcus* species (Appendix 1).^13^ We defined probable pathogens as known pathogens including but not limited to *Enterobacterales* (e.g., *E coli*, *Salmonella* Paratyphi A, Paratyphi B, Paratyphi C, *S.* Typhi, non-typhoidal serovars of *S. enterica*), *Enterococcus* species, Group A *Streptococcus*, *Haemophilus influenzae*, *Neisseria meningitidis*, *Staphylococcus aureus*, and *Streptococcus pneumoniae*. If one blood culture yielded multiple probable pathogen isolates, it was defined as ‘mixed probable pathogens.’ If a participant had two or more blood cultures with probable pathogens within 14 days apart, we counted only the first positive blood culture.

### Metrics of sentinel bloodstream infections

As in our earlier study,^4^ we calculated the following metrics of the bloodstream infections from prevalence studies: (i) prevalence of *S.* Typhi among all probable pathogen isolates as done by Mackenzie et al. for pneumococcal disease prevalence,^14^ (ii) rank order of *S.* Typhi among probable pathogen isolates, (iii) ratio of *S.* Typhi prevalence to *E. coli* prevalence, and (iv) ratio of *S.* Typhi prevalence to ‘stably endemic organisms’ defined as *E. coli, S. aureus*, or *S. pneumoniae*. Rank order was calculated only if *S.* Typhi was isolated at least once. For the latter two metrics, we calculated ratios for only *E. coli*, or if one of *E. coli, S. aureus*, or *S. pneumoniae* were isolated at least once.

### Data analysis

We excluded study sites with <10 bloodstream isolates classified as probable pathogens. First, we described the article characteristics including study design. We then described each bloodstream infection metric across study sites, stratified by hospitalisation status. Next, we analysed each bloodstream infection metric across study sites, stratifying by hospitalisation status to account for potential variations in pathogen epidemiology between inpatient and outpatient settings. The association between prevalence metrics and typhoid incidence estimate was assessed using meta-regression. To achieve a symmetrical distribution for analysis, typhoid incidence estimates were log-transformed using the general inverse variance method ^15^. For sites reporting zero typhoid incidence, a continuity correction of 0.01 cases per 100,000 person-years was applied prior to transformation.^16^ We used the inverse of person-years of observation as the standard error. We tested the linearity of each metric using restricted cubic splines. Logarithmic transformations were applied to both the *S.* Typhi to *E. coli* ratio and the *S.* Typhi to “stably endemic organisms” ratio, with outliers truncated at the 0.99 percentile. To account for uncertainty in the typhoid incidence estimates, meta-regression analyses were repeated using the lower and upper bounds of the reported 95% CI or uncertainty ranges.

We categorised the abstracted typhoid incidence estimate at each study site into low, (<10/100,000 person-years), medium, (10-100/100,000 person-years), and high (>100/100,000 person-years).^17^ In univariate ordinal regression analysis, we assessed the association of each of the metrics of bloodstream infections with typhoid incidence level. In a sensitivity analysis, we repeated ordinal regression analysis while omitting study sites with outliers exceeding the 0.99 percentile for either the prevalence metrics or the incidence estimates. Additionally, we performed subgroup analyses to evaluate age-specific incidence estimates. We considered a p-value below 0.05 as statistically significant.

#### Model derivation

We derived the final model using study sites with complete data for the prevalence metrics. We explored combinations of metrics to identify the optimal model for predicting typhoid incidence level based on the likelihood ratio test or the lowest Akaike Information Criterion (AIC). We selected the final model based on model performance and parsimony to ensure ease of implementation.

#### Model performance

The model generated three outcomes: (1) low typhoid incidence versus medium or high typhoid incidence, (2) medium typhoid incidence versus low or high typhoid incidence, and (3) high typhoid incidence versus low or medium typhoid incidence. To assess the internal validity, we performed a bootstrap validation with 500 resamples.^18^ We compared the performance of this bootstrap model with the performance of the original model. The average difference between these performances across all iterations was defined as the optimism. We reported the discriminative value using the concordance statistic (C-statistic) for each outcome.^19^ Additionally, model calibration was assessed using the optimism-corrected intercept and slope to measure the agreement between predicted risks and observed outcomes.^20^

#### Decision curve analysis and diagnostic accuracy

To determine the practical value of the model in a public health context, we performed a decision curve analysis (DCA).^21^ While traditional diagnostic measures like sensitivity and specificity quantify model accuracy, DCA evaluates the *net benefit* of applying the model across a range of threshold probabilities. The net benefit balances the benefit of correctly identifying a high incidence site (‘true positive’) against the harm or cost of unnecessary intervention at a low-incidence site (‘false positive’). In this context, the threshold probability represents the risk level at which a policymaker justifies an intervention (e.g., a vaccination campaign). For instance, a cautious policymaker with a low threshold might trigger a vaccination campaign based on even a small predicted probability of high typhoid incidence. The resulting decision curve compares our model against two default scenarios: intervening at all sites or intervening at none. Additionally, we assessed diagnostic accuracy measures, including sensitivity and specificity, across a range of cut-offs for site-specific predicted probabilities. We simulated the predicted probabilities of the model for a range of values of the metrics of sentinel bloodstream infections.

#### Extrapolation

Beyond *S.* Typhi, we assessed the transportability of the prevalence metrics to estimate the incidence of non-typhoidal *Salmonella* (NTS) and *Salmonella* Paratyphi A. From the included articles, we abstracted the incidence of NTS or *Salmonella* Paratyphi A when available. In the aforementioned metrics, we replaced *S.* Typhi with non-typhoidal *Salmonella* or *Salmonella* Paratyphi A, respectively. We repeated the same meta-regression and univariate ordinal regression analysis. All analyses were performed in R version 4.4.1 (packages: epiR, MASS, meta, rms).

### Ethics statement

This study was a secondary analysis of published data and existing de-identified data and was approved by University of Otago Human Ethics Committee HD23/012. Local ethical approval of the original studies covered the use for de-identified data for the purpose of this study.^5,6,9–11,22–24^

### Role of the funding source

The funder of the study had no role in study design, data collection, data analysis, data interpretation, or writing of the report.

## Results

Of 12 articles identified from the updated systematic review (Appendix 2), we included data from eight articles. ^6,8–11,22,24,25^ In addition, we added one article identified by consultation with experts published before 1 January 2018 reporting on the Typhoid Fever Surveillance in Africa Program.^5^ The nine articles yielded data on 29 eligible study sites. Of the nine articles, three articles reported typhoid incidence estimated based on sentinel surveillance with multipliers, three articles were vaccine trials using Typbar-TCV (Bharat Biotech International, Hyderabad, India) in the intervention arm, one article used active household-based surveillance, and one article used sentinel surveillance with multipliers and active household-based surveillance (Table 1). Data collection ranged from 1 March 2010 through 9 April 2021.

**Table 1.**
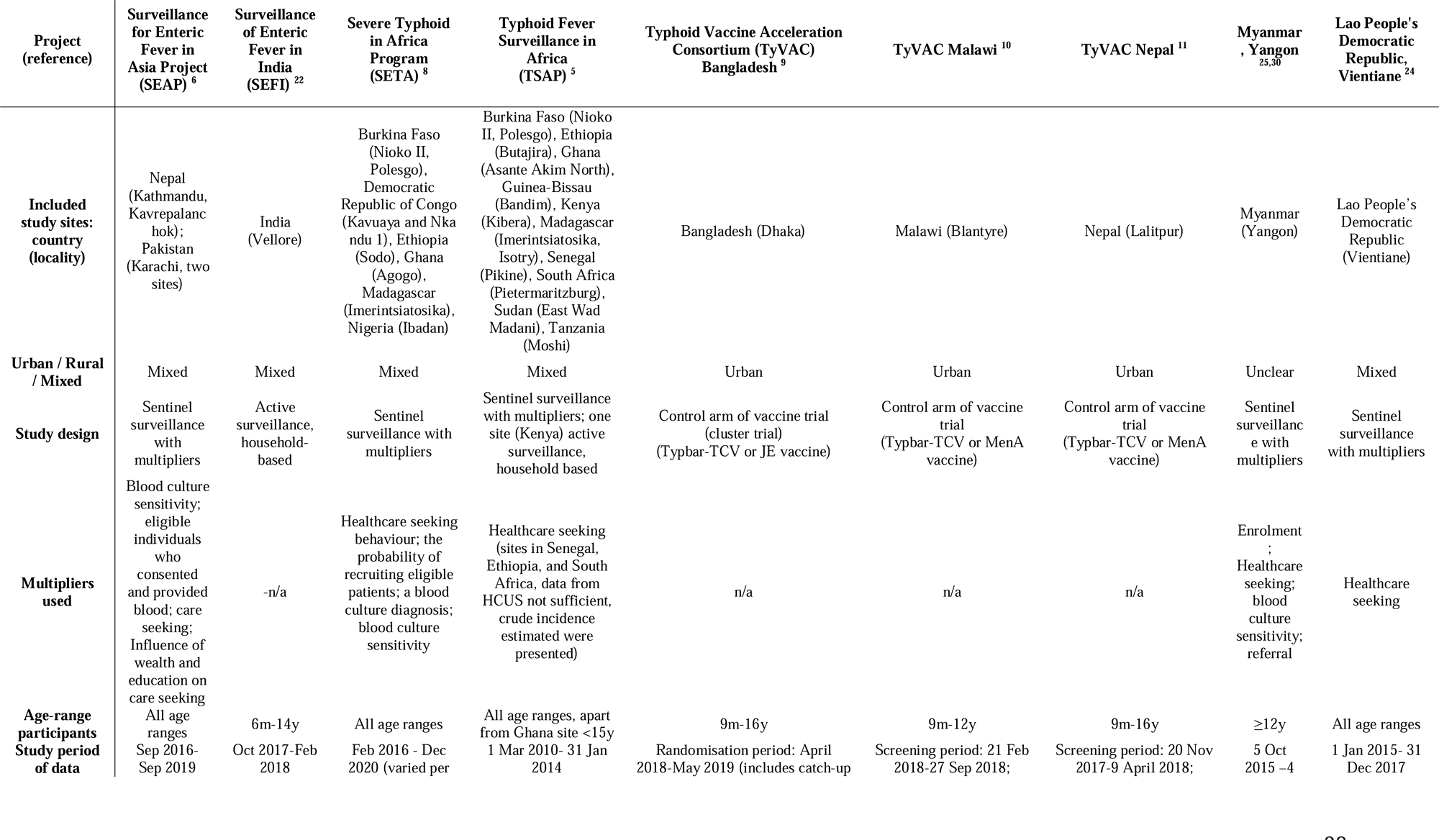
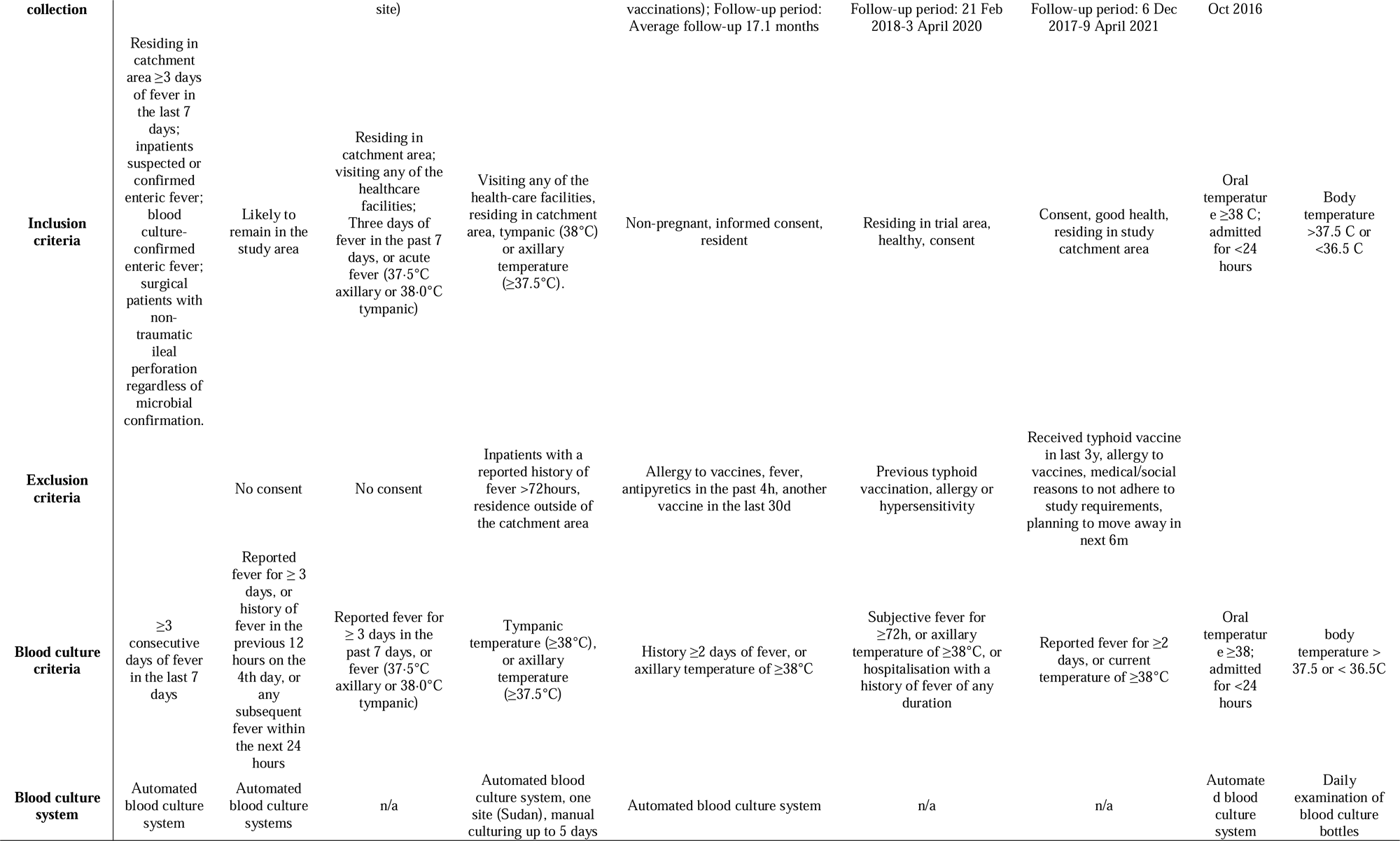
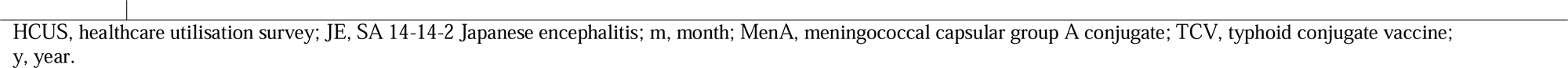
Characteristics of included articles that reported typhoid incidence estimates and available prevalence data of bloodstream infections from sentinel sites, published 2017-2024.

The fever duration to be eligible for blood culture collection was ≥3 days in three articles, ≥3 days or fever measured at presentation in one article, ≥2 days of fever or fever measured at presentation in two articles, or measured fever of any duration at presentation in three articles. Of 29 study sites, 20 (68.9%) reported data from fourteen countries of the United Nations (UN) African region, and nine (31.0%) reported data from six countries of the UN Asian region (Appendix 3). Across study sites, the total number of blood cultures was 70,886 with a median (IQR) of 1,550 (976-4,239) blood cultures per study site. Across 28 study sites with available data, the median (IQR) proportion of blood cultures with any growth, including likely contaminants, was 10.0% (5.7-14.2%) and the median (IQR) proportion of blood cultures yielding a probable pathogen was 3.6% (2.7-6.1%). Of 29 study sites, this resulted in data of 4,625 probable pathogen isolates. Across study sites, the number of blood cultures was not correlated with the proportion of blood cultures with probable pathogen isolates.

### Metrics of sentinel bloodstream infections

For prevalence of *S.* Typhi isolates among all probable pathogens across 29 study sites, the median (IQR) prevalence was 39.8% (19.1-73.7%). For rank order of *S.* Typhi among all probable pathogens across 28 study sites, the median (IQR) rank was 1 (1-2) (Appendix 5). For the ratio of *S.* Typhi to *E. coli* across 26 study sites, the median (IQR) ratio was 4.9 (1.3-13.3), and for the ratio *S.* Typhi to ‘stably endemic organisms’ across 29 study sites, the median (IQR) ratio was 1.3 (0.5-10.7). The metrics of sentinel bloodstream infections did not differ by hospitalisation status for seven study sites with available data (Appendix 6).

### Typhoid incidence estimates

The reported typhoid incidence estimate (95% CI) by study site ranged from 0 (0-0) per 100,000 person-years to 1,189 (490-2,940) per 100,000 person-years, with a median (IQR) of 140 (28-319) per 100,000 person-years (Figure 1). Of 29 study sites, five (17.2%) were classified as low typhoid incidence sites, six (20.7%) as medium, and 18 (62.1%) as high (Table 2). Ten (90.9%) of the eleven study sites classified as low typhoid incidence and medium typhoid incidence were from the African region. Of the 18 study sites classified as high typhoid incidence, ten (55.6%) were from the African region and eight (44.4%) were from the Asian region. Compared with the assigned level of the typhoid incidence estimate, the lower bound of the 95% CI was in a lower typhoid incidence level for four study sites, and the upper bound of the 95% CI was in a higher typhoid incidence level in three study sites.

**Figure 1:**
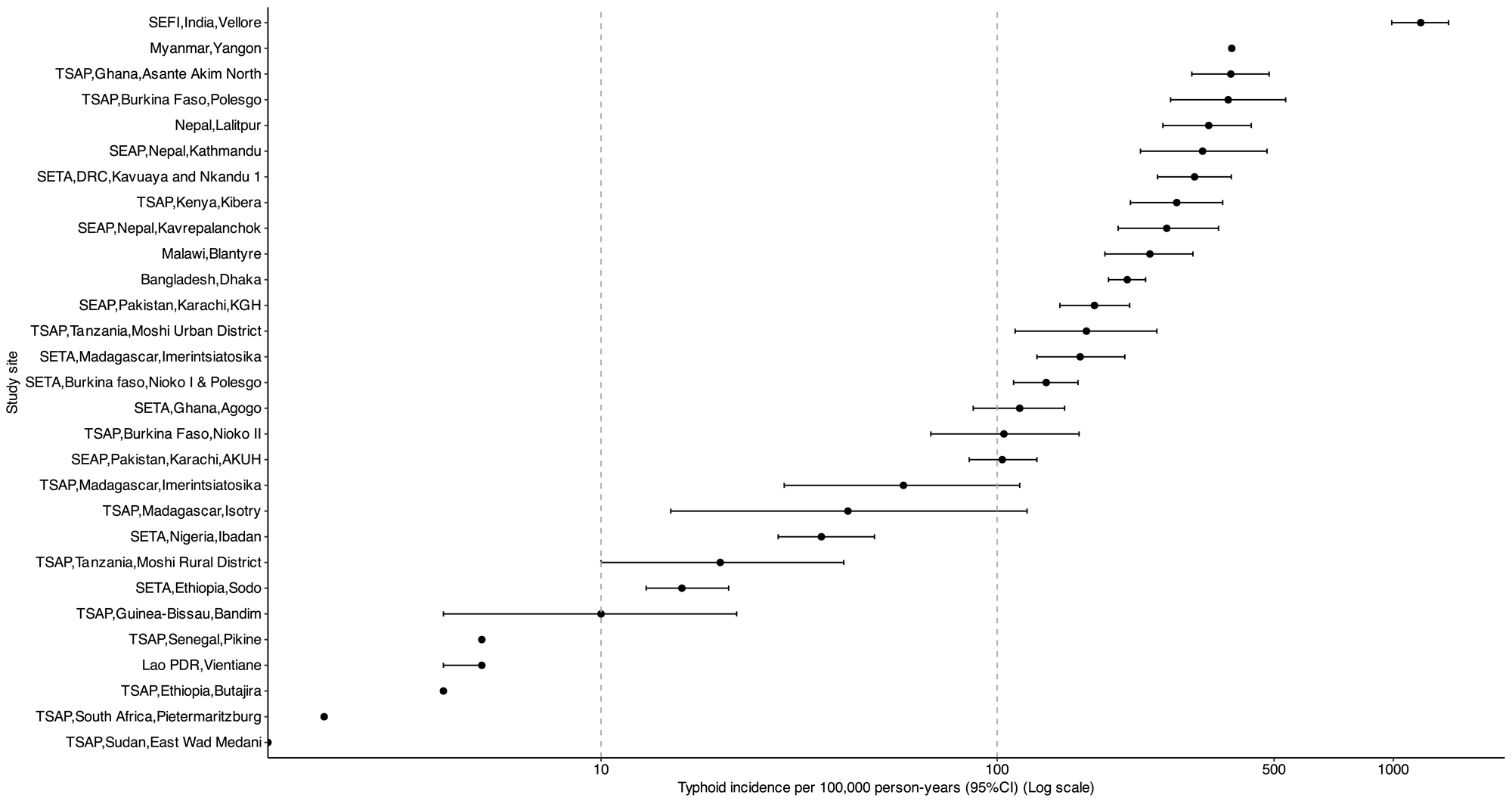
Distribution of the typhoid incidence estimates across study sites, 2017-2024 Legend: AKUH, Aga Khan University Hospital; CI, confidence interval; KGH, Kharadar General Hospital; NA, not available; SEAP, Surveillance for Enteric Fever in Asia Project; SEFI, Surveillance of Enteric Fever in India; SETA, Severe Typhoid Fever Surveillance in Africa; TSAP, Typhoid Fever Surveillance in Africa Program

**Table 2.**
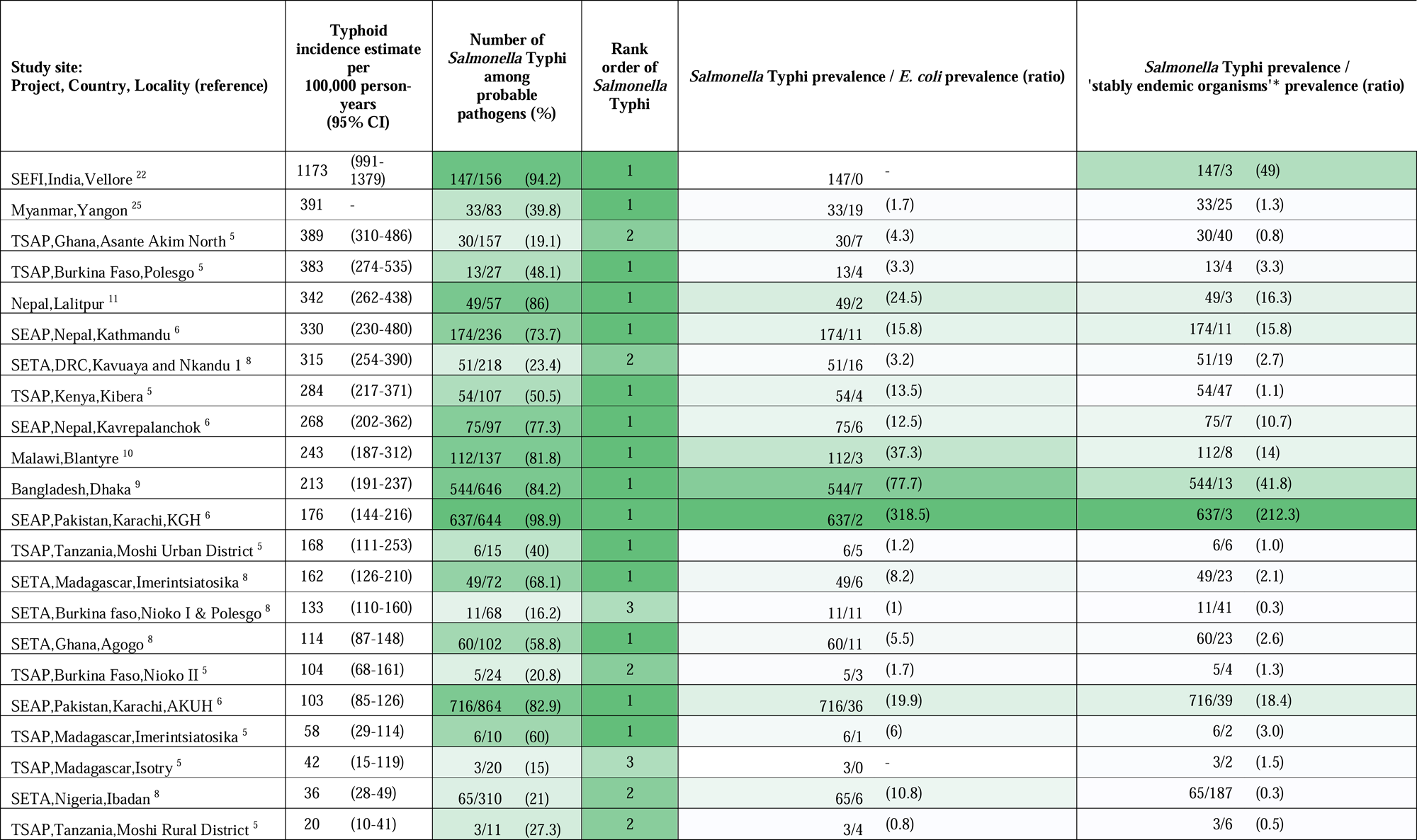
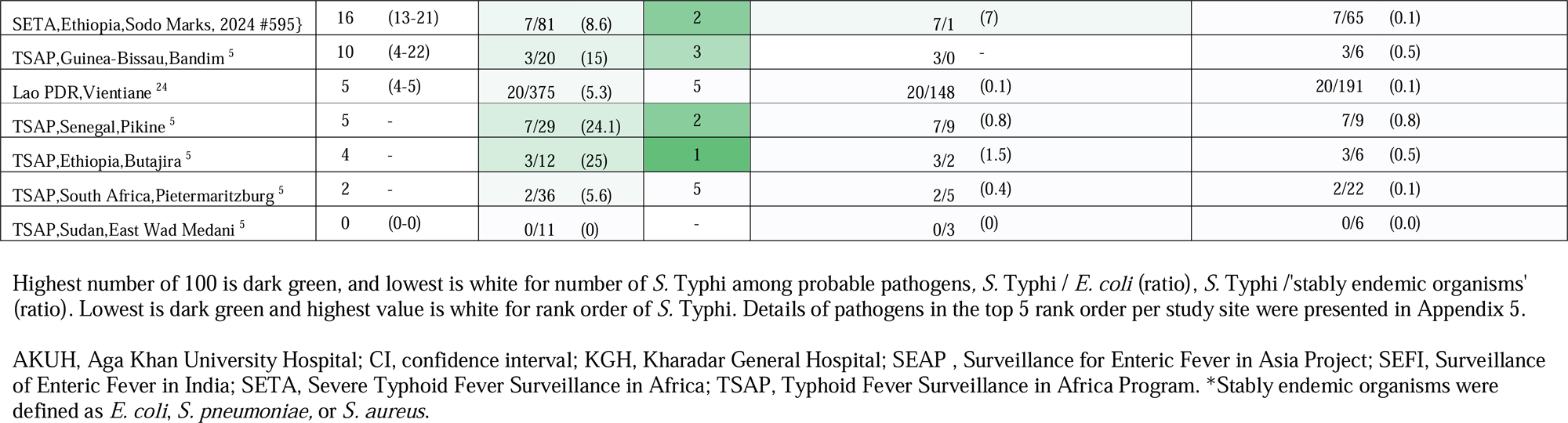
Reported typhoid incidence estimates and metrics from bloodstream infections from sentinel sites, sorted by typhoid incidence estimate, 2017-2024.

In the meta-regression, all metrics of sentinel bloodstream infections were associated with log typhoid incidence. The beta (95%CI) was 0.05 (0.03-0.07) for each 1% increase in prevalence of *S.* Typhi among all probable pathogens, −0.97 (−1.39 - −0.55) for rank order of *S.* Typhi among all probable pathogens, 0.86 (0.62-1.10) for log ratio of *S.* Typhi to *E. coli*, and 0.84 (0.62-1.06) for the log ratio of *S.* Typhi to ‘stably endemic organisms’ (Appendix 7, Appendix 8). For the association of the metrics of sentinel bloodstream infections with the lower and upper bounds of the typhoid incidence estimate, similar directions for the associations were observed.

In univariate ordinal regression analysis, all metrics of sentinel bloodstream infections were associated with typhoid incidence level. The OR (95%CI) was 1.08 (1.04-1.17) for each 1% increase in prevalence of *S.* Typhi among all probable pathogens, 0.24 (0.07-0.55) for rank order of *S.* Typhi among all probable pathogens, 2.91 (1.49-7.43) for the log ratio of *S.* Typhi to *E. coli*, and 3.75 (1.81-10.66) for the log ratio of *S.* Typhi to ‘stably endemic organisms’ (Appendix 7). In sensitivity analyses with the exclusion of sites with outliers in Vellore, India,^22^ (typhoid incidence estimate) and Karachi, Pakistan^6^ (prevalence of *S.* Typhi, ratio *S.* Typhi to *E. coli*, ratio *S.* Typhi to ‘stably endemic organisms’) similar results for the association with typhoid incidence level were observed. Subgroup analyses for age were planned, but due to the limited sample size not feasible.

### Model performance

We compared models for typhoid incidence level with various combinations of prevalence metrics in 25 study sites with complete data (Appendix 9). Although the model with the log ratio of *S.* Typhi to ‘stably endemic organisms’ as sole predictor had better performance compared with the model with prevalence of *S.* Typhi of probable pathogens as sole predictor, the latter was chosen as final model. This decision was based on parsimony and ease of implementation. The c-statistic of this model was 0.86 and the optimism-corrected c-statistic (95%CI) was 0.86 (0.75-0.99). The slope and intercept of the optimism-corrected calibration were 0.02 and 0.93, respectively. For the 29 study sites with available data, model predictions were calculated for typhoid incidence level. The c-statistic (95%CI) of the predicted probabilities was 0.87 (0.58-0.97) for the identification of low typhoid incidence sites, 0.76 (0.51-0.91) for medium typhoid incidence sites, and 0.88 (0.69-0.96) for high typhoid incidence sites. For low, medium, and high typhoid incidence levels, apparent calibration showed intercepts of 0.00, −0.03, and 0.03 and slopes of 1.47, 0.76, and 0.94, respectively (Appendi× 10). The decision curve analysis showed that the model for low typhoid incidence and medium typhoid incidence did not have a higher net benefit compared with intervention for all or none of the study sites (Appendix 11). For the outcome of high typhoid incidence, the model showed a higher net benefit compared with the intervention for all or none of the study sites.

For low typhoid incidence, the predicted probability cutoffs of <10% and <50% had sensitivity (95% CI) of 0.00 (0.00-0.52) and 0.40 (0.05-0.85), respectively, and specificity (95% CI) of 0.00 (0.00-0.14) and 0.38 (0.19-0.50), respectively (Table 3). For medium typhoid incidence, the predicted probability cutoffs of >10% and >50% had sensitivity (95%) of 0.33 (0.04-0.78) and 0.83 (0.36-1.00), respectively, and specificity (95%CI) of 0.43 (0.23-0.66) and 1.00 (0.85-1.00), respectively. For high typhoid incidence, the predicted probability cut-offs of >10% and >50% had sensitivity (95% CI) of 0.78 (0.52-0.94) and 1.00 (0.81-1.00), respectively, and specificity (95% CI) of 0.09 (0.00-0.41) and 0.91 (0.59-1.00), respectively.

**Table 3.**
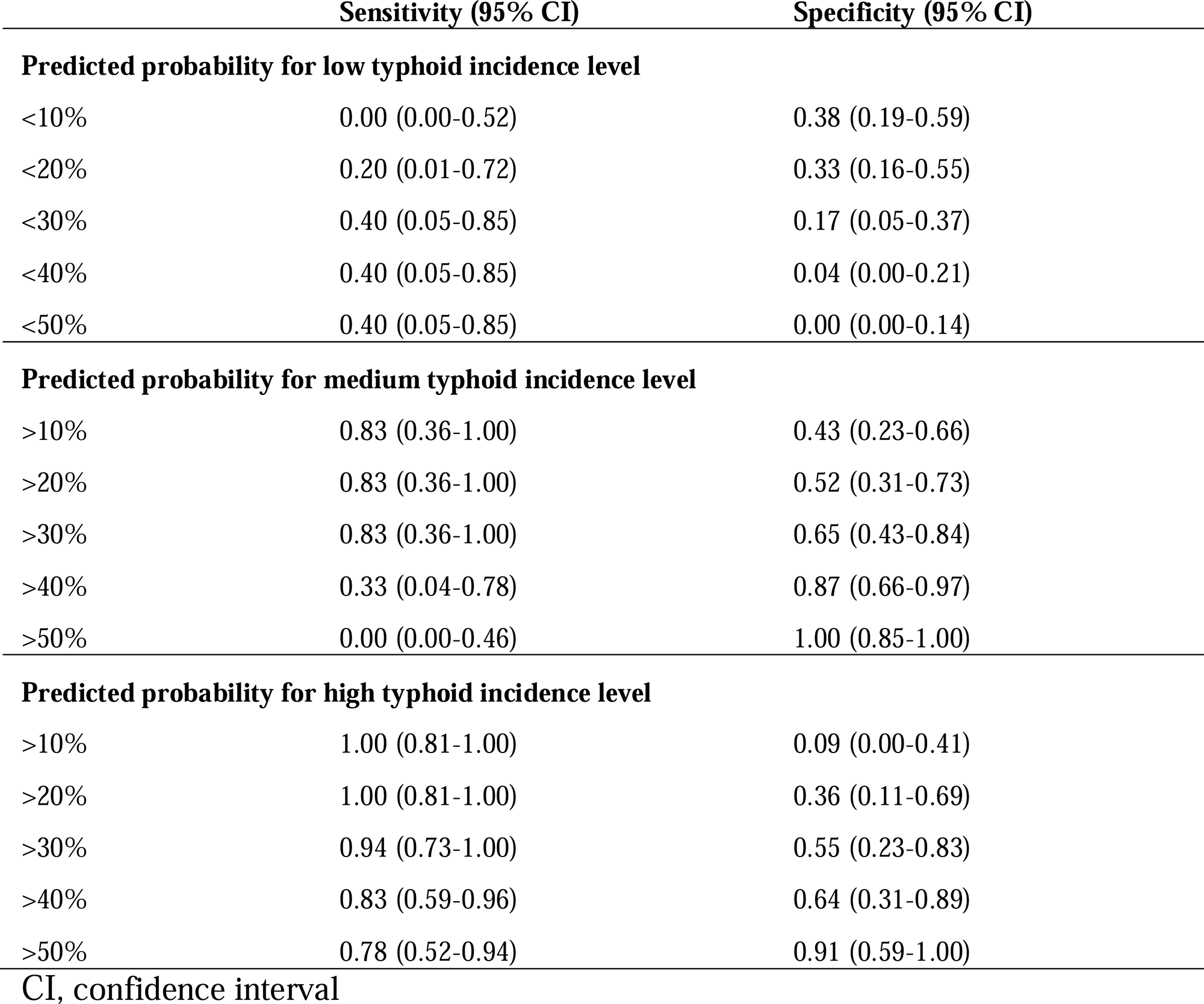
Diagnostic value for cut-offs of predicted probabilities for typhoid incidence levels according to the final model, 2017-2024.

Simulations of model predictions of low, medium, and high typhoid incidence for the range of prevalence of *S.* Typhi values are shown in Figure 2 and Appendix 12.

**Figure 2:**
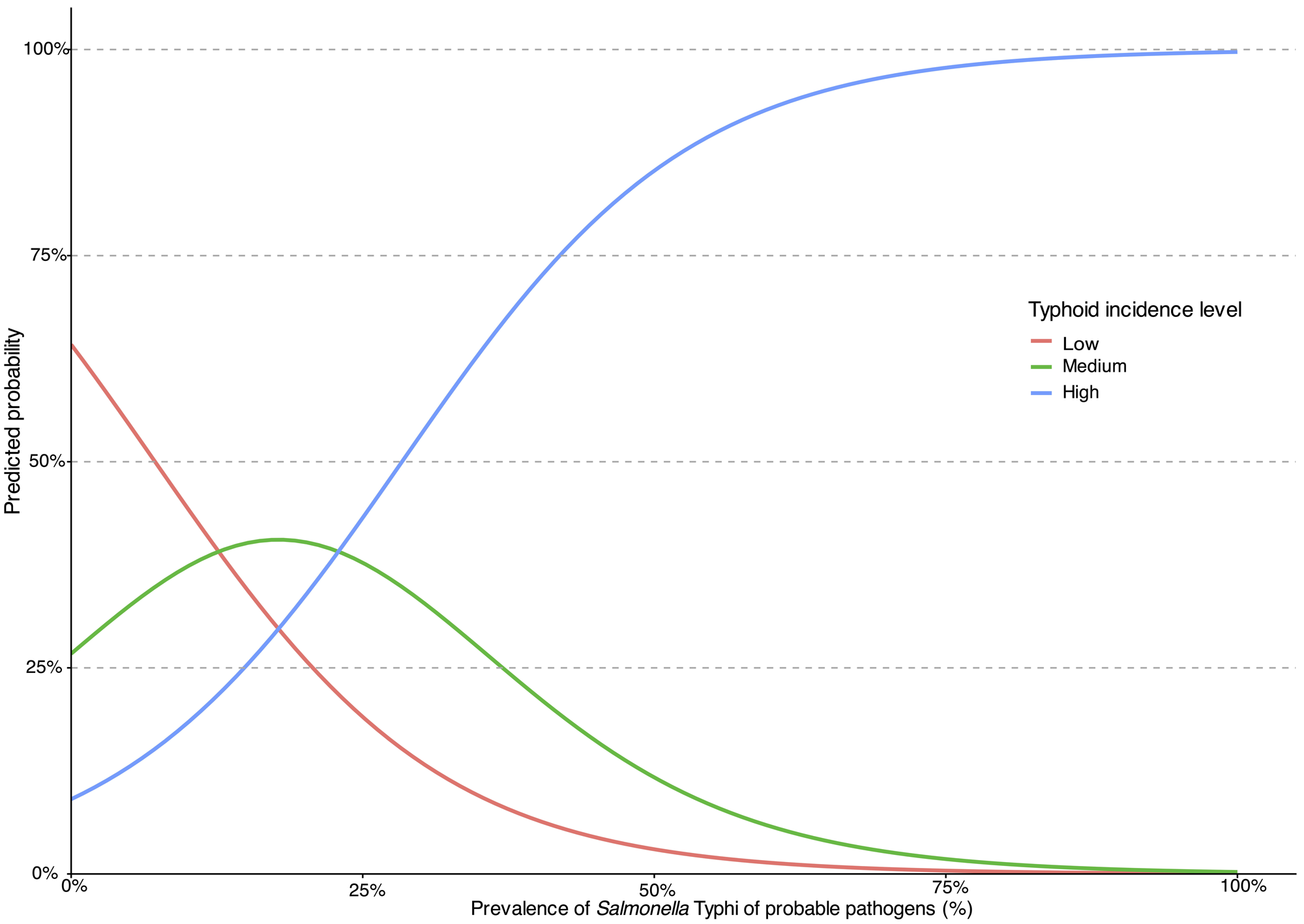
Predicted probabilities for typhoid incidence levels according to the model based on prevalence of S. Typhi of probable pathogens Legend: Predicted probabilities (y-axis) for typhoid incidence level: low (red), medium (green), and high (blue) based on the prevalence of S. Typhi of probable pathogens (x-axis). Lines represent the predicted probabilities based on prevalence of S. Typhi according to the ordinal regression model.

In the extrapolation for non-typhoidal salmonella, associations of the prevalence metrics with incidence estimates were similar for 13 African study sites (Appendix 13). For extrapolation to *Salmonella* Paratyphi A, data were available for seven study sites and were insufficient for analysis.

## Discussion

In this study, we examined the relationship of *S.* Typhi incidence with metrics from sentinel community-onset bloodstream infections and developed a pragmatic tool for policy decisions for control of typhoid fever. Our model, based on prevalence of *S.* Typhi among probable pathogen isolates, demonstrated robust discriminative ability, as evidenced by the c-statistic, and achieved good calibration when predicting high incidence. Notably, the model’s diagnostic value was sensitive to the chosen probability cut-off. For example, a 30% probability cut-off for high typhoid incidence yielded high sensitivity, suggesting it could effectively assist policymakers in identifying sites with low-to-medium typhoid incidence.

Unlike previous typhoid incidence estimation efforts that relied on country-level covariates and complex disease burden modelling,^26,27^ our approach proposes a pragmatic tool based on localised prevalence metrics which relies on diagnostic laboratories capable of processing blood cultures and identifying pathogens. This design allows the tool to be implemented in local settings without the need for complex transformations or extensive country covariate datasets, increasing its practical applicability. Although more easily available than large prospective surveillance studies, obtaining blood culture data in lower resource settings remains challenging due to systemic barriers in resources, infrastructure and technical expertise.

Several factors may influence the better discrimination of high typhoid incidence. Because our data were derived from population-based surveillance and vaccine trials, the study sites may be inherently biased toward high incidence locations. Indeed, more than half of the included sites were classified as having high typhoid incidence, which likely contributed to the model’s stronger performance in those sites compared to medium- or low-incidence sites. In addition, substantial heterogeneity was observed across sites with respect to country context, eligible age ranges, intensity of surveillance, blood culture collection criteria, and the spectrum of identified pathogens (Appendix 5). These differences should be considered when interpreting the results and may limit generalisability to settings with a different casemix or thresholds for blood culture collection. Last, typhoid incidence may vary over time and by location. Therefore, metrics of bloodstream infection should ideally be calculated over multiple years to provide a reliable indication of underlying typhoid incidence.

The extrapolation of our framework to non-typhoidal salmonella confirmed the association between prevalence metrics and incidence estimates. Data from *Salmonella* Paratyphi A incidence, however, were limited, precluding a validation for this disease in our study. These findings suggest that while sentinel metrics are promising proxies for the incidence of typhoid and non-typhoidal salmonella, further exploration and validation across other diseases is required.

Policy decisions on typhoid vaccine introduction and investments in non-vaccine prevention and control strategies are based on diverse forms of evidence, including local disease incidence, cost-effectiveness, and access to safe water and improved sanitation.^28,29^ Simple metrics, such as the prevalence of *S.* Typhi among sentinel community-onset bloodstream infections, tend to be more widely available than typhoid incidence estimates from prospective population-based or hybrid surveillance studies. As such, our approach can provide insights on typhoid fever incidence when contemporary incidence estimates are unavailable.

Strengths of our study included the use of contemporaneous, isolate-level data from typhoid incidence studies identified through an updated systematic review. Second, we developed a prediction model based on simple bloodstream infection metrics that are readily available and can be easily calculated. Third, instead of expert opinion, we employed a data-driven approach to model development. Our study had some limitations. The number of included study sites was limited. However, the underlying data encompassed a large population under surveillance and included 4,625 probable pathogen isolates. To maximise the number of study sites, we identified studies from our previous systematic review and undertook repeated data requests from original study authors. Nevertheless, developing a model using a limited number of sites introduces the risk of overfitting. To mitigate this, we performed internal validation using resampling techniques to address optimism. Owing to the small sample size and the nature of the available data, internal–external or fully external validation was not feasible.

Where direct typhoid incidence estimates are unavailable, we provide a practical approach to support policy decisions by identifying settings with low, medium, or high typhoid incidence using routinely collected bloodstream infections data from sentinel sites. This approach may help inform prioritisation of typhoid conjugate vaccine introduction and complementary investments in water, sanitation, and hygiene.

## Author contributions

Conceptualisation: NNH, SM, CSM, JAC

Data curation: NNH, SM, CSM, FA, JA, BB, ASC, SD, IFD, KD, DOG, JJ, HJ, JJ, FK, JL, XL, FM, PN, SRN, KN, PDP, AJP, FQ, FQ, TR, JCS, MS, SS, BTT, DT, MV, MV, MTY, JAC

Formal analysis: NNH

Funding acquisition: JAC

Methodology: NNH, SM, CSM, JW, JAC

Project administration: JAC

Supervision: JAC

Validation: JAC

Visualization: NNH, JW

Writing original draft preparation: NNH

Writing review and editing: SM, CSM, JW, FA, JA, BB, ASC, SD, IFD, KD, DOG, JJ, HJ, JJ, FK, JL, XL, FM, SRN, KN, PN, PDP, AJP, FQ, FQ, TR, JCS, MS, SS, BTT, DT, MV, MV, MTY, JAC

All authors reviewed and approved this manuscript before submission, had access to the data, and have final responsibility for the decision to submit for publication.

## Data availability statement

The underlying data have been deposited in Dataverse: https://doi.org/10.7910/DVN/JTTFYL. This includes the underlying data to replicate all analysis and the supplementary material with detailed methods and results. Local ethical approvals did not include open posting of the isolate-level data. De-anonymised data regarding this study can be requested from the corresponding author. Proposals should be submitted by the study team via the corresponding author by email.

## Notes

**Conflicts of interest:** The authors declare no competing financial interests or personal relationships that could have appeared to influence the work reported in this paper.

### Competing Interest Statement

The authors have declared no competing interest.

### Funding Statement

This study was funded by Gates Foundation (GF) grant INV-030857.

### Author Declarations

This study was a secondary analysis of published data and existing de-identified data and was approved by University of Otago Human Ethics Committee HD23/012.

