## Supplementary material for "Predicting *Salmonella* Typhi incidence using prevalence metrics from sentinel studies of community-onset bloodstream infections"

### **Appendix 1 – Definition of contaminants**

**Definition of contaminants**

*Bacillus* species

Coagulase-negative *Staphylococcus*

*Corynebacterium* species (diphteroids)

Micrococcus species

*Cutibacterium* (previously *Propionibacterium*)

Alpha-haemolytic *Streptococcus* (other than *Strep. pneumoniae*)

*Pseudomonas* (other than *P. aeruginosa*)

Other environmental non-fermenting gram-negative rods

### **Appendix 2 – Eligible articles reporting on typhoid incidence**

| **Citation** | **First**  **author** | **Publication**  **year** | **Title** | **Project** | **Notes** |
| --- | --- | --- | --- | --- | --- |
| ^1^ | Chanthavilay | 2020 | Estimation of Incidence of Typhoid and Paratyphoid Fever in Vientiane, Lao People’s Democratic Republic |  | Included |
| ^2^ | Cutting | 2022 | Facility-based disease surveillance and Bayesian hierarchical modeling to estimate endemic typhoid fever incidence, Kilimanjaro Region, Tanzania, 2007–2018 |  | Not included because overlap with TSAP |
| ^3^ | Garrett | 2022 | Incidence of typhoid and paratyphoid fever in Bangladesh, Nepal, and Pakistan: results of the Surveillance for Enteric Fever in Asia Project | Surveillance for Enteric Fever in Asia Project  (SEAP) | Data from study sites Pakistan and Nepal are included |
| ^4^ | John | 2018 | Estimating the incidence of enteric fever in children in India: a multi-site, active fever surveillance of pediatric cohorts | Surveillance of Enteric Fever in India  (SEFI) | Data from study site Vellore included |
| ^5^ | Marks | 2022 | The Severe Typhoid in Africa Program: Incidences of Typhoid Fever in Burkina Faso, Democratic Republic of Congo, Ethiopia, Ghana, Madagascar, and Nigeria | Severe Typhoid in Africa Program  (SETA) | Included: 5 study sites were not eligible as no typhoid incidence was reported. One study site Madagascar, Mahajanga was excluded because only <10 probable pathogens were isolated. |
| ^6^ | Marks | 2017 | Incidence of invasive salmonella disease in sub-Saharan Africa: a multicentre population-based surveillance study | Typhoid Fever Surveillance in Africa  (TSAP) | Included |
| ^7^ | Meiring | 2021 | Burden of enteric fever at three urban sites in Africa and Asia: a multicentre population-based study | Strategic Typhoid Alliance across Africa and Asia (STRATAA) | Not included |
| ^8^ | Ng’eno | 2023 | Dynamic Incidence of Typhoid Fever over a 10-Year Period (2010-2019) in Kibera, an Urban Informal Settlement in Nairobi, Kenya |  | Not included because overlap with TSAP |
| ^9,10^ | Oo | 2019 | Incidence of Typhoid and Paratyphoid Fevers Among Adolescents and Adults in Yangon, Myanmar |  | Included, prevalence data  abstracted from publication ^10^ |
| ^11^ | Patel | 2021 | Safety and Efficacy of a Typhoid Conjugate Vaccine in Malawian Children | Typhoid Vaccine Acceleration Consortium (TyVAC) - Malawi | Data from the control arm included |
| ^12^ | Qadri | 2021 | Protection by vaccination of children against typhoid fever with a Vi-tetanus toxoid conjugate vaccine in urban Bangladesh: a cluster-randomised trial | Typhoid Vaccine Acceleration Consortium (TyVAC) - Bangladesh | Data from the cluster control arm  are included |
| ^13^ | Shakya | 2021 | Efficacy of typhoid conjugate vaccine in Nepal: final results of a phase 3, randomised, controlled trial | Typhoid Vaccine Acceleration Consortium (TyVAC) - Nepal | Data from the control arm included |
| ^14^ | Yousafzai | 2021 | Effectiveness of typhoid conjugate vaccine against culture confirmed *Salmonella* enterica serotype Typhi in an extensively drug-resistant outbreak setting of Hyderabad, Pakistan: a cohort study |  | Not included. |

### **Appendix 3 – Map of countries of the study sites of typhoid incidence and prevalence of community-onset bloodstream infections, published 2017-2024**

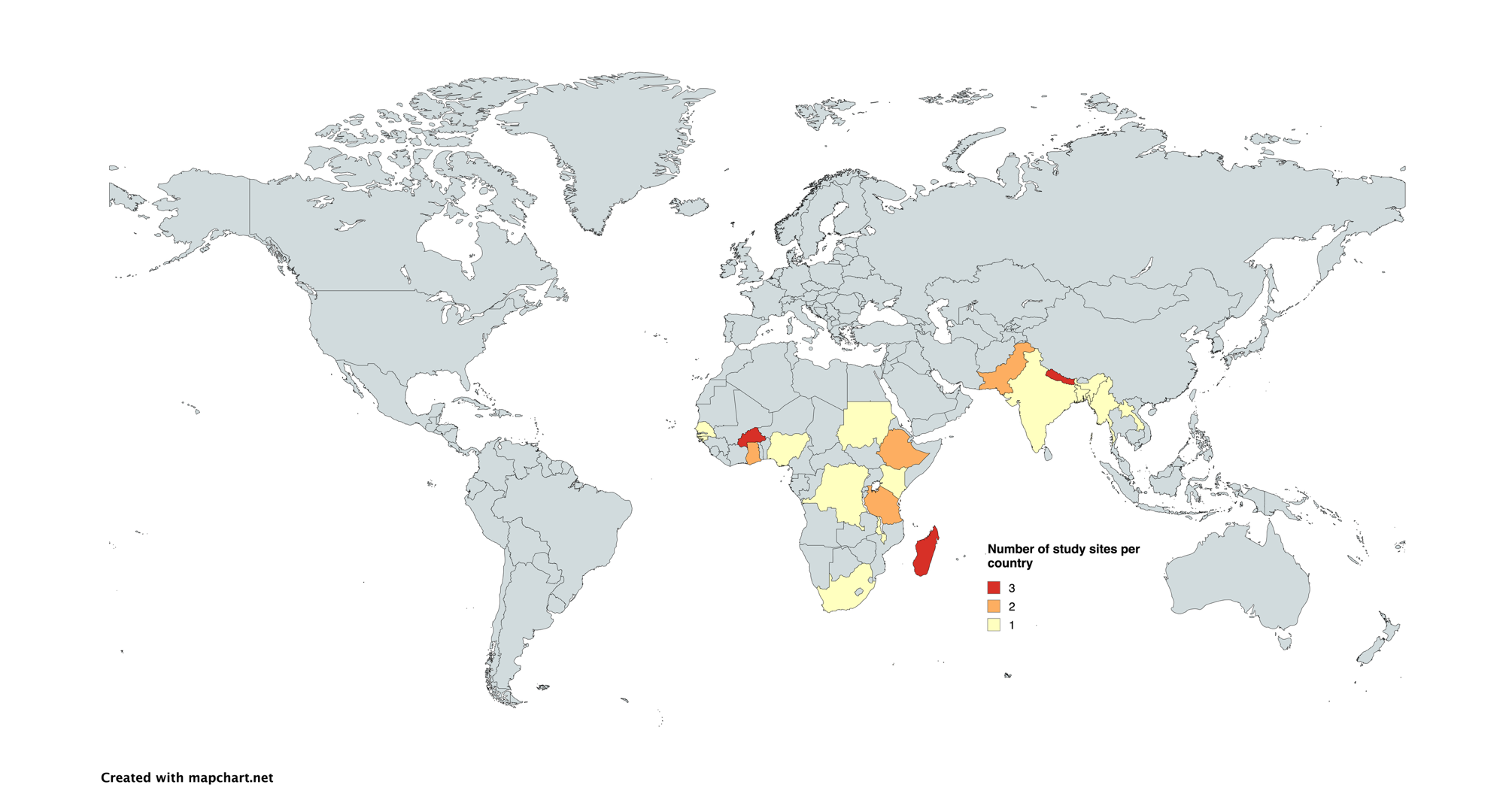

### **Appendix 4 – Distribution of blood culture results across study sites, sorted from high typhoid incidence to low typhoid incidence study sites**

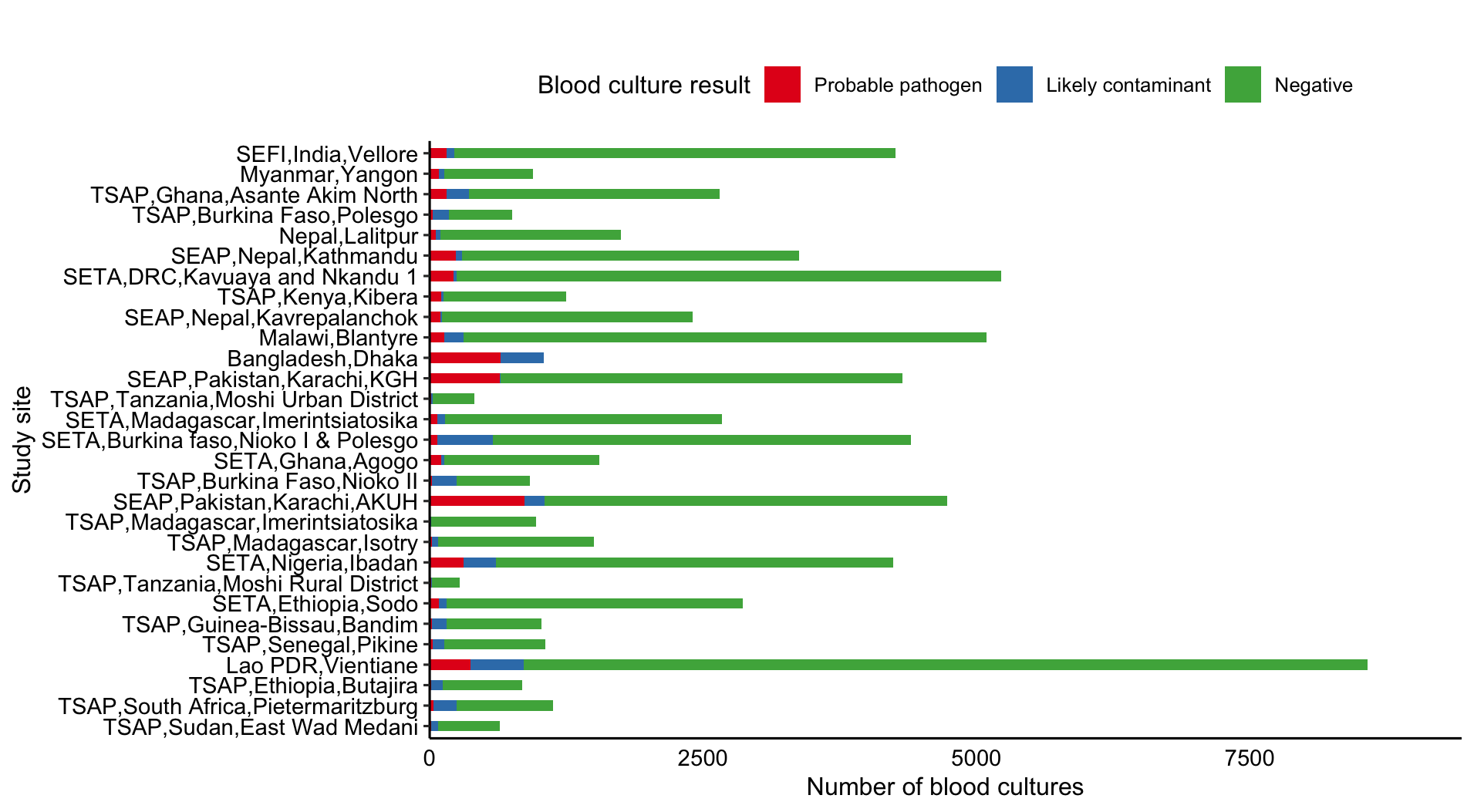

SEAP, Surveillance for Enteric Fever in Asia Project (SEAP); SEFI, Surveillance of Enteric Fever in India; SETA, Severe Typhoid in Africa Program (SETA); TSAP, Typhoid Fever Surveillance in Africa.

Blood culture negative data were not available for Bangladesh, Dhaka.

### **Appendix 5 – Top five rank order probable pathogens isolated from bloodstream infections across study sites, sorted by highest typhoid incidence study site**

| **Study site (Project, Country, Locality)** | **Reference** | **Number of probable pathogens** | **Top five rank order of probable pathogens: Isolate genus species (number of isolates)** | | | |  |
| --- | --- | --- | --- | --- | --- | --- | --- |
|  |  |  | **1** | **2** | **3** | **4** | **5** |
| Bangladesh,Dhaka | ^12^ | 646 | *S.* Typhi (544) | *S.* Paratyphi A (73) | *E. coli* (7) | *Salmonella* nontyphoidal serovars (5) | *Staphylococcus*  *aureus* (4) |
| Lao PDR,Vientiane | ^1^ | 375 | *E. coli* (148) | *Klebsiella*  *pneumoniae* (49) | *Staphylococcus*  *aureus* (34) | *Salmonella* nontyphoidal serovars (23) | *S.* Typhi (20) |
| Malawi,Blantyre | ^11^ | 137 | *S.* Typhi (112) | *Salmonella* nontyphoidal serovars (12) | *Streptococcus*  *pneumoniae* (4) | *E. coli* (3) | *Cryptococcus*  *neoformans* (1) |
| Myanmar,Yangon | ^9^ | 83 | *S.* Typhi (33) | *E. coli* (19) | *S.* Paratyphi A (10) | *Klebsiella*  *pneumoniae* (6) | *Staphylococcus*  *aureus* (6) |
| Nepal,Lalitpur | ^13^ | 57 | *S.* Typhi (49) | *S.* Paratyphi A (4) | *E. coli* (2) | *Enterococcus* species (1) | *Staphylococcus*  *aureus* (1) |
| SEAP,Nepal,Kathmandu | ^3^ | 236 | *S.* Typhi (174) | *S.* Paratyphi A (42) | *E. coli* (11) | *Klebsiella*  *pneumoniae* (5) | *Aeromonas* species (1) |
| SEAP,Nepal,Kavrepalanchok | ^3^ | 97 | *S.* Typhi (75) | *S.* Paratyphi A (13) | *E. coli* (6) | *Enterococcus* species (1) | *Salmonella* nontyphoidal serovars (1) |
| SEAP,Pakistan,Karachi,AKUH | ^3^ | 864 | *S.* Typhi (716) | *S.* Paratyphi A (87) | *E. coli* (36) | *Salmonella* nontyphoidal serovars (4) | *Klebsiella*  *pneumoniae* (3) |
| SEAP,Pakistan,Karachi,KGH | ^3^ | 644 | *S.* Typhi (637) | *E. coli* (2) | *S.* Paratyphi A (2) | *Enterobacter* species (1) | *Salmonella* nontyphoidal serovars (1) |
| SEFI,India,Vellore | ^4^ | 156 | *S.* Typhi (147) | *Salmonella* nontyphoidal serovars (2) | *S.* Paratyphi A (2) | *Streptococcus*  *pneumoniae* (2) | *Enterococcus* species (1) |
| SETA,Burkina Faso,  Nioko I & Polesgo | ^5^ | 68 | Staphylococcus  aureus (29) | *E. coli* (11) | *S.* Typhi (11) | *Salmonella* nontyphoidal serovars (9) | *Citrobacter freundii* (2) |
| SETA,DRC,  Kavuaya and Nkandu 1 | ^5^ | 218 | *Salmonella* nontyphoidal serovars (144) | *S.* Typhi (51) | *E. coli* (16) | *Staphylococcus*  *aureus* (3) | *Enterococcus* species (2) |
| SETA,Ethiopia,Sodo | ^5^ | 81 | Staphylococcus  aureus (63) | *S.* Typhi (7) | *Pseudomonas*  *aeruginosa* (4) | *Enterobacter*  *cloacae* (2) | *Erwinia* species (2) |
| SETA,Ghana,Agogo | ^5^ | 102 | *S.* Typhi (60) | *Salmonella* nontyphoidal serovars (13) | *E. coli* (11) | *Staphylococcus*  *aureus* (6) | *Streptococcus*  *pneumoniae* (6) |
| SETA,Madagascar,  Imerintsiatosika | ^5^ | 72 | *S.* Typhi (49) | Streptococcus  pneumoniae (14) | *E. coli* (6) | *Staphylococcus*  *aureus* (3) | *NA* |
| SETA,Nigeria,Ibadan | ^5^ | 310 | Staphylococcus  aureus (180) | *S.* Typhi (65) | *Salmonella* nontyphoidal serovars (28) | *E. coli* (6) | *Candida albicans* (4) |
| TSAP,Burkina Faso,Nioko II | ^6^ | 24 | *Salmonella* nontyphoidal serovars (10) | *S.* Typhi (5) | *E. coli* (3) | *Enterococcus*  faecalis (2) | *Aeromonas* species (1) |
| TSAP,Burkina Faso,Polesgo | ^6^ | 27 | *S.* Typhi (13) | *E. coli* (4) | *Salmonella* nontyphoidal serovars (4) | *Proteus mirabilis* (2) | *Pseudomonas*  *aeruginosa* (2) |
| TSAP,Ethiopia,Butajira | ^6^ | 12 | *S.* Typhi (3) | *E. coli* (2) | *Leclercia*  *adecarboxylata* (2) | *Staphylococcus*  *aureus* (2) | *Streptococcus*  *pneumoniae* (2) |
| TSAP,Ghana,  Asante Akim North | ^6^ | 157 | *Salmonella* nontyphoidal serovars (59) | *S.* Typhi (30) | *Klebsiella*  *pneumoniae* (22) | *Streptococcus*  *pneumoniae* (17) | *Staphylococcus*  *aureus* (16) |
| TSAP,Guinea-Bissau,Bandim | ^6^ | 20 | *Salmonella* nontyphoidal serovars (8) | *Staphylococcus*  *aureus* (4) | *S.* Typhi (3) | *Enterobacter*  *cloacae* (2) | *Streptococcus*  *pneumoniae* (2) |
| TSAP,Kenya,Kibera | ^6^ | 107 | *S.* Typhi (54) | *Staphylococcus*  *aureus* (33) | *Streptococcus*  *pneumoniae* (10) | *Salmonella* nontyphoidal serovars (6) | *E. coli* (4) |
| TSAP,Madagascar,  Imerintsiatosika | ^6^ | 10 | *S.* Typhi (6) | *E. coli* (1) | *Salmonella* nontyphoidal serovars (1) | *Streptococcus*  *pneumoniae* (1) | *Yersinia pestis* (1) |
| TSAP,Madagascar,Isotry | ^6^ | 20 | *Candida* species (6) | *Enterobacter*  *cloacae* (3) | *S.* Typhi (3) | *Klebsiella oxytoca* (2) | *Enterobacter*  *asburiae* (1) |
| TSAP,Senegal,Pikine | ^6^ | 29 | *E. coli* (9) | *S.* Typhi (7) | *Salmonella* nontyphoidal serovars (4) | *S.* Paratyphi A (3) | *Enterobacter*  *cloacae* (2) |
| TSAP,South Africa,  Pietermaritzburg | ^6^ | 36 | *Staphylococcus*  *aureus* (10) | *Streptococcus*  *pneumoniae* (7) | *E. coli* (5) | *Cryptococcus*  *neoformans* (4) | *S.* Typhi (2) |
| TSAP,Sudan,  East Wad Medani | ^6^ | 11 | *E. coli* (3) | *Staphylococcus*  *aureus* (3) | *Proteus*  *mirabilis* (2) | *Enterococcus*  *casseliflavus* (1) | *Pasteurella*  *pneumotropica* (1) |
| TSAP,Tanzania,  Moshi Rural District | ^6^ | 11 | *E. coli* (4) | *S.* Typhi (3) | *Clostridium*  species (1) | *Salmonella* nontyphoidal serovars (1) | *Staphylococcus*  *aureus* (1) |
| TSAP,Tanzania,  Moshi Urban District | ^6^ | 15 | *S.* Typhi (6) | *E. coli* (5) | *Proteus*  *mirabilis* (1) | *Salmonella* nontyphoidal serovars (1) | *Streptococcus*  *pneumoniae* (1) |

SEAP, Surveillance for Enteric Fever in Asia Project (SEAP); SEFI, Surveillance of Enteric Fever in India; SETA, Severe Typhoid in Africa Program (SETA); TSAP, Typhoid Fever Surveillance in Africa.

### **Appendix 6 - Metrics from sentinel bloodstream infection stratified by hospitalisation status, 2017-2024**

| **Study site** | **Hospital admission** | **Number of *S.* Typhi among probable pathogens (%)** | **Rank order of *S.* Typhi** | ***S.* Typhi prevalence / *E. coli* prevalence (ratio)** | ***S.* Typhi prevalence /**  **'stably endemic organisms'* prevalence (ratio)** |
| --- | --- | --- | --- | --- | --- |
| Bangladesh,Dhaka ^12^ | no | 522/621 (84.1) | 1 | 522/7 (74.6) | 522/13 (40.2) |
| Bangladesh,Dhaka ^12^ | yes | 22/25 (88.0) | 1 |  |  |
| Lao PDR,Vientiane ^1^ | yes | 20/375 (5.3) | 5 | 20/148 (0.1) | 20/191 (0.1) |
| Malawi,Blantyre ^11^ | yes | 12/14 (85.7) | 1 |  |  |
| SETA,Burkina faso,Nioko I & Polesgo Marks, 2024 #595} | yes | 1/1 (100) | 1 |  |  |
| SETA,DRC,Kavuaya and Nkandu 1 ^5^ | yes | 10/22 (45.5) | 1 | 10/6 (1.7) | 10/6 (1.7) |
| SETA,Ethiopia,Sodo ^5^ | yes | 1/13 (7.7) | 4 |  | 1/10 (0.1) |
| SETA,Ghana,Agogo ^5^ | yes | 21/40 (52.5) | 1 | 21/2 (10.5) | 21/9 (2.3) |
| SETA,Nigeria,Ibadan ^5^ | yes | 26/118 (22) | 2 | 26/5 (5.2) | 26/57 (0.5) |
| TSAP,Burkina Faso,Nioko II ^6^ | no | 5/23 (21.7) | 2 | 5/3 (1.7) | 5/4 (1.3) |
| TSAP,Burkina Faso,Nioko II ^6^ | yes | 0/1 (0) |  |  |  |
| TSAP,Burkina Faso,Polesgo ^6^ | no | 13/27 (48.1) | 1 | 13/4 (3.3) | 13/4 (3.3) |
| TSAP,Ethiopia,Butajira ^6^ | no | 3/10 (30) | 1 | 3/2 (1.5) | 3/5 (0.6) |
| TSAP,Ethiopia,Butajira ^6^ | yes | 0/2 (0) |  |  | 0/1 (0) |
| TSAP,Ghana,Asante Akim North ^6^ | yes | 30/157 (19.1) | 2 | 30/7 (4.3) | 30/40 (0.8) |
| TSAP,Guinea-Bissau,Bandim ^6^ | no | 2/15 (13.3) | 3 |  | 2/5 (0.4) |
| TSAP,Guinea-Bissau,Bandim ^6^ | yes | 1/5 (20.0) | 3 |  | 1/1 (1) |
| TSAP,Kenya,Kibera ^6^ | no | 54/107 (50.5) | 1 | 54/4 (13.5) | 54/47 (1.1) |
| TSAP,Madagascar,Imerintsiatosika ^6^ | no | 6/10 (60.0) | 1 | 6/1 (6) | 6/2 (3) |
| TSAP,Madagascar,Isotry ^6^ | no | 3/20 (15.0) | 3 |  | 3/2 (1.5) |
| TSAP,Senegal,Pikine ^6^ | no | 5/15 (33.3) | 2 | 5/6 (0.8) | 5/6 (0.8) |
| TSAP,Senegal,Pikine ^6^ | yes | 2/14 (14.3) | 4 | 2/3 (0.7) | 2/3 (0.7) |
| TSAP,South Africa,Pietermaritzburg ^6^ | yes | 2/36 (5.6) | 5 | 2/5 (0.4) | 2/22 (0.1) |
| TSAP,Sudan,East Wad Medani ^6^ | no | 0/11 (0.0) |  | 0/3 (0) | 0/6 (0) |
| TSAP,Tanzania,Moshi Rural District ^6^ | no | 1/4 (25.0) | 3 | 1/1 (1) | 1/2 (0.5) |
| TSAP,Tanzania,Moshi Rural District ^6^ | yes | 2/7 (28.6) | 2 | 2/3 (0.7) | 2/4 (0.5) |
| TSAP,Tanzania,Moshi Urban District ^6^ | no | 2/5 (40.0) | 1 |  | 2/1 (2) |
| TSAP,Tanzania,Moshi Urban District ^6^ | yes | 4/10 (40.0) | 2 | 4/5 (0.8) | 4/5 (0.8) |

CI, confidence interval; NA, not available; SETA, Severe Typhoid Fever Surveillance in Africa; TSAP, Typhoid Fever Surveillance in Africa Program. *Stably endemic organisms were defined as *E. coli*, *S. pneumoniae,* or *S. aureus.*

### **Appendix 7 – Association of metrics from bloodstream infections from sentinel sites with typhoid incidence by meta-regression and univariate ordinal regression analysis, 2017-2024**

|  | **Meta-regression for log typhoid incidence estimate** | **Univariate ordinal regression analysis for low, medium and high typhoid incidence estimate levels^±^** |
| --- | --- | --- |
| **Metrics of sentinel bloodstream infection** | **Beta (95%CI)** | **OR (95%CI)** |
| Each 1% increase in prevalence of *S.* Typhi among probable pathogens (n=29) | 0.05 (0.03-0.07) | 1.08 (1.04-1.17) |
| Each increase in rank order of *S.* Typhi among probable pathogens (n=28) | -0.97 (-1.39- -0.55) | 0.24 (0.07-0.55) |
| Log *S.* Typhi to *E. coli* ratio (n=25) | 0.86 (0.62-1.10) | 2.91 (1.49-7.43) |
| Log *S.* Typhi to 'stably endemic organisms' ratio *(n=29) | 0.84 (0.62-1.06) | 3.75 (1.81-10.7) |

OR, odds ratio

± Low typhoid incidence (<10/100,000 person-years); medium typhoid incidence (10-100/100,000 person-years); high typhoid incidence (>100/100,000 person-years)

*Stably endemic organisms were defined as *E. coli*, *S. pneumoniae,* or *S. aureus.*

### **Appendix 8 – Bubble plots of meta-regression of metrics from sentinel blood stream infections with log typhoid incidence (A, prevalence of *S.* Typhi among probable pathogens; B, rank order of *S.* Typhi among probable pathogens; C, ratio log *S.* Typhi to *E. coli* ratio; D, log ratio *S.* Typhi to ‘stably endemic organisms’ ratio)**

A:

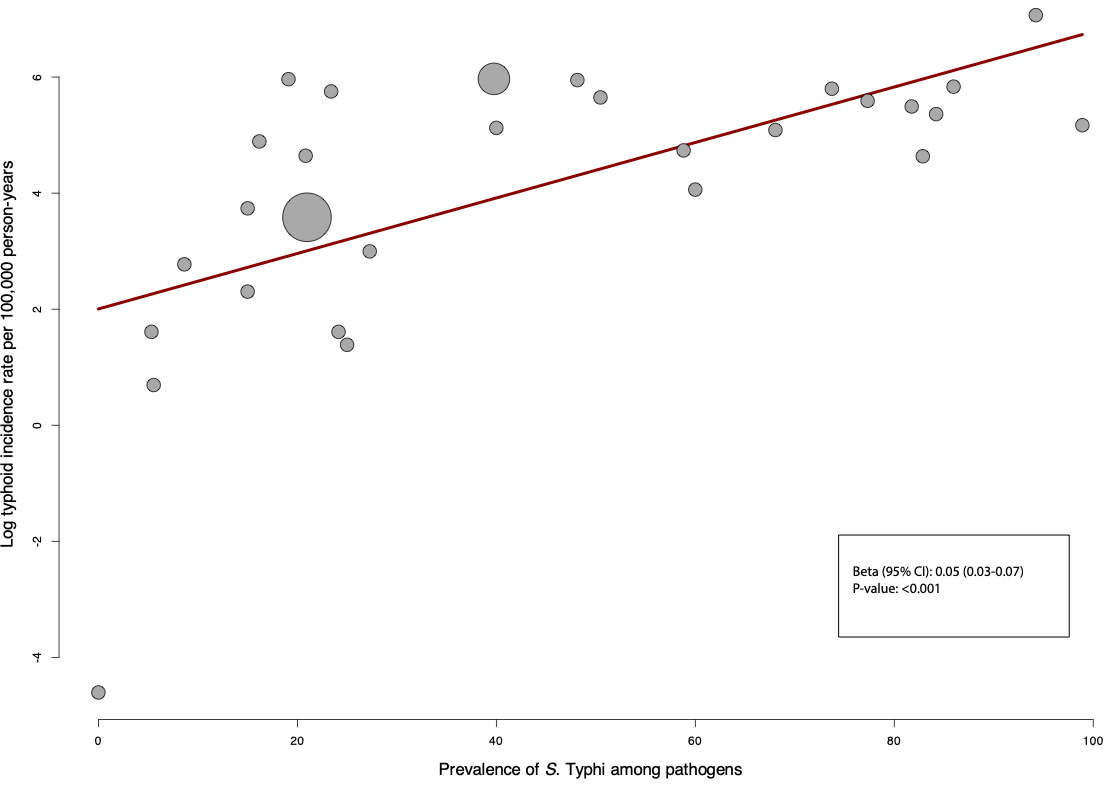

B:

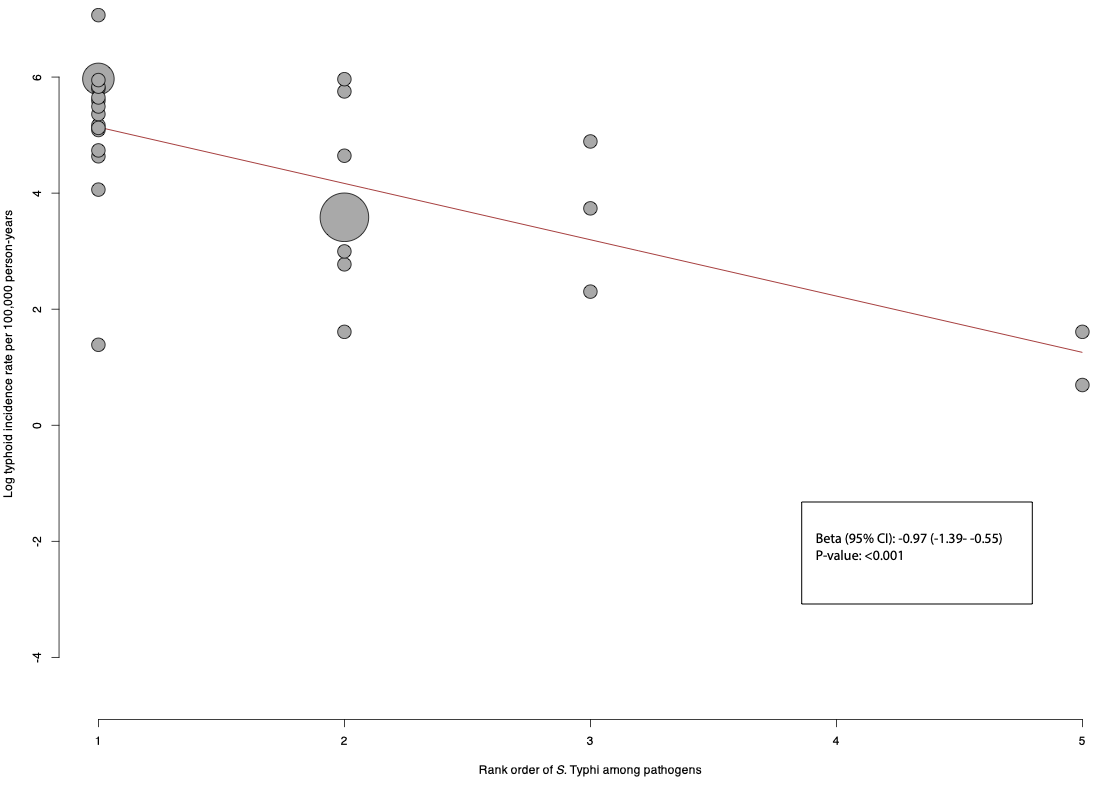

C:

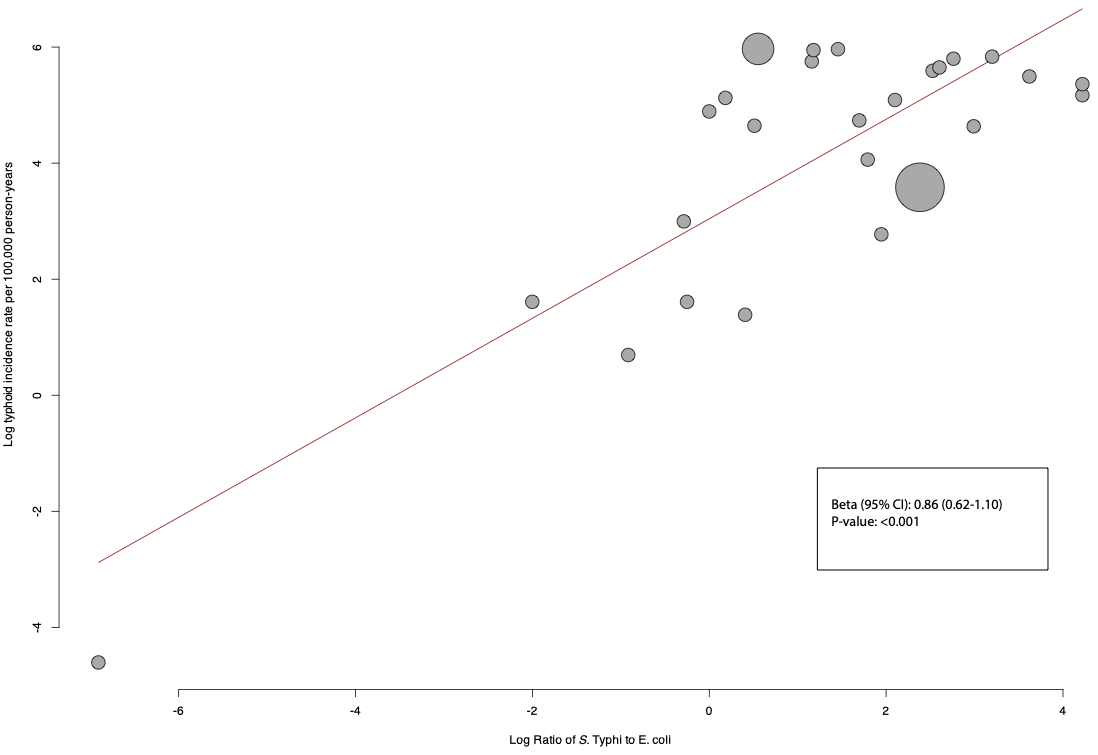

D:

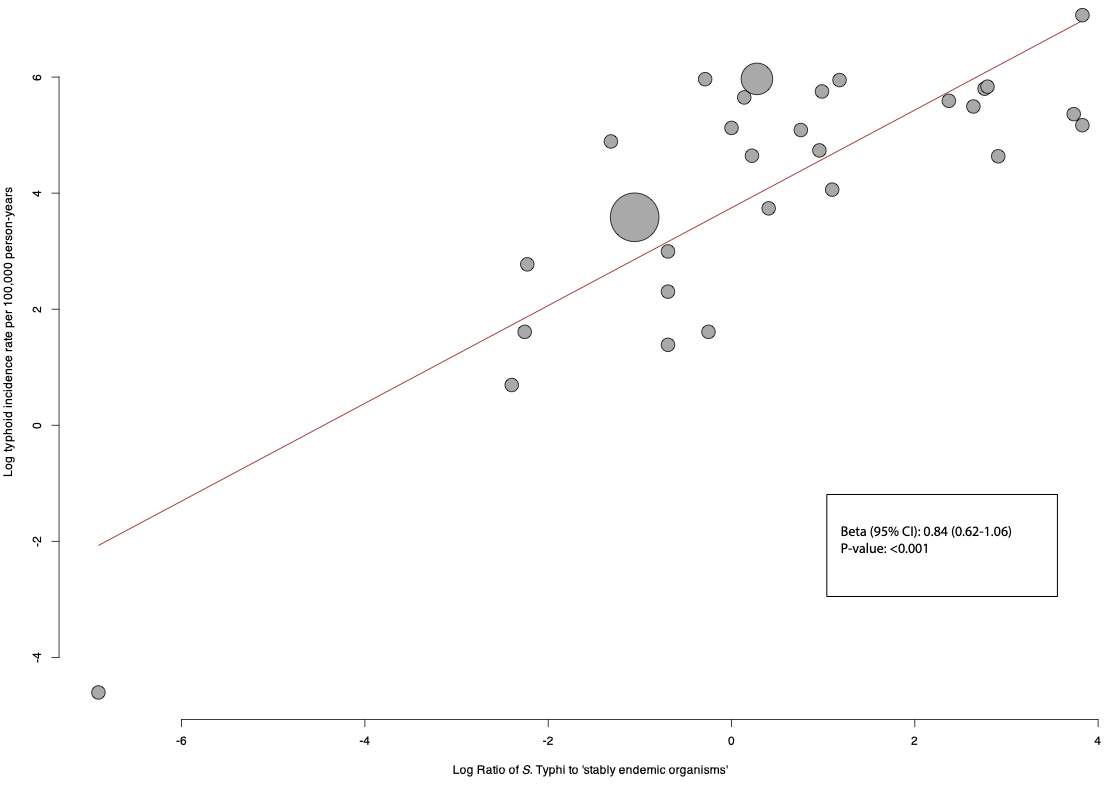

### **Appendix 9 – Summary of model comparisons for metrics of bloodstream infections of sentinel sites for typhoid incidence level in ordinal regression analysis, 2017-2024**

Models were compared using the likelihood ratio test and the AIC. Model selection was based on model performance, parsimony, and ease of implementation.

Model 1	Prevalence of *S.* Typhi of probable pathogens

Model 2	Rank order of *S.* Typhi

Model 2b Prevalence of *S.* Typhi of probable pathogens and rank order of *S.* Typhi

Model 3	Log ratio of *S.* Typhi to *E. coli*

Model 3b Prevalence of *S.* Typhi of probable pathogens and log ratio of *S.* Typhi to *E. coli*

Model 4	Log ratio of *S.* Typhi to 'stably endemic organisms'

Model 4b Prevalence of Typhi of probable pathogens and log ratio of *S.* Typhi to 'stably endemic organisms'

|  | **P-value** | **AIC** | **Decision** |
| --- | --- | --- | --- |
| Model 1 vs Model 2 | 0.33 | 37.4 vs 39.0 | Keep model 1 |
| Model 1 vs Model 2b | 1 | 37.4 vs 38.5 | Keep model 1 |
| Model 1 vs Model 3 | 1 | 37.4 vs 38.5 | Keep model 1 |
| Model 1 vs Model 3b | 0.16 | 37.4 vs 37.4 | Keep model 1 based on parsimony |
| Model 1 vs Model 4 | <0.01 | 37.4 vs 34.5 | Keep model 4 |
| Model 4 vs Model 4b | 0.85 | 34.5 vs 36.4 | Keep model 4 |

AIC, Akaike Information Criterion

Final model: model 1 (prevalence of *S.* Typhi of probable pathogens as sole predictor) was selected based on parsimony and ease of calculation, since calculation of ratios and log transformation is a complicating factor for policy makers*.*

The formula to calculate the predicted probabilities is presented below:

$P (Incidence) = low) = \frac{1}{1+e^{-(LP1\text{)}}}$

$$P (Incidence) = medium) = \frac{1}{1+e^{-(LP2\text{)}}}-\frac{1}{1+e^{-(LP1\text{)}}}$$

$$P (Incidence) = high) = 1- \frac{1}{1+e^{-(LP2\text{)}}}$$

*LP* refers to the linear predictor in a proportional odds logistic regression model. The LPs for the thresholds are defined as follows:

*LP_1_* (low vs*.* medium or high): *LP_1_* = 0.5846 (Intercept) - 0.0813 * prevalence of *S.* Typhi of pathogens

*LP_2_* (low/medium vs high): *LP_2_* =2.3051 (Intercept) - 0.0813 * prevalence of *S.* Typhi of pathogens

*LP_3_* (high vs medium/low): *LP_3=_* -2.3051 (Intercept) + prevalence of *S.* Typhi of pathogens

### **Appendix 10 – Calibration plots: predicted probability vs observed proportion of the model for typhoid incidence level, 2017-2024**

##### Low typhoid incidence vs medium and high typhoid incidence

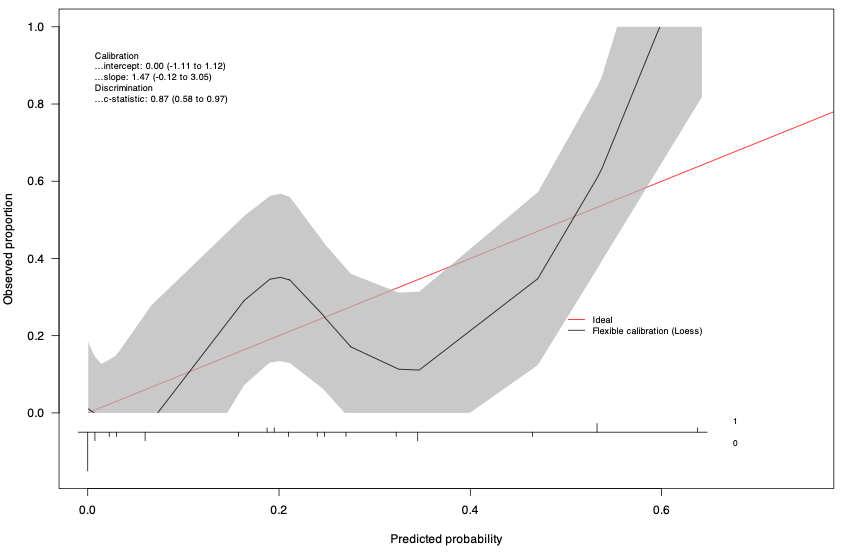

##### Medium typhoid incidence vs low and high typhoid incidence

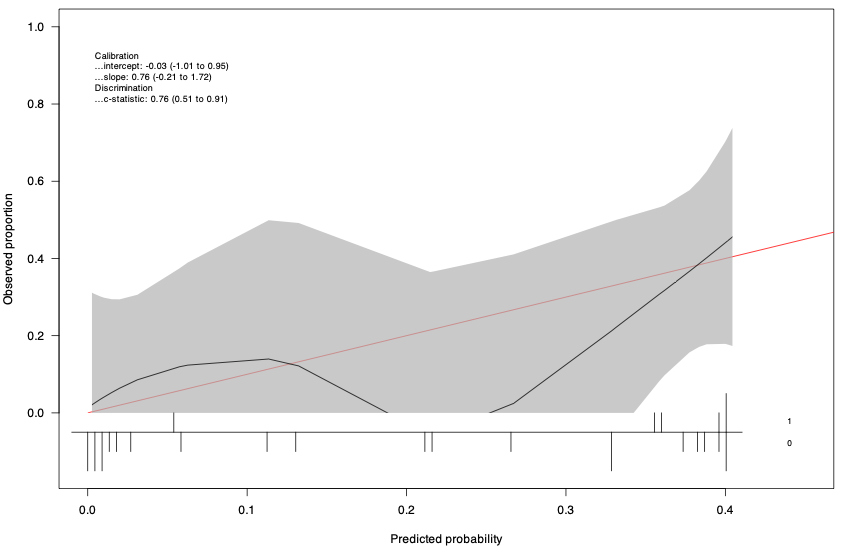

##### High typhoid incidence vs low and medium typhoid incidence

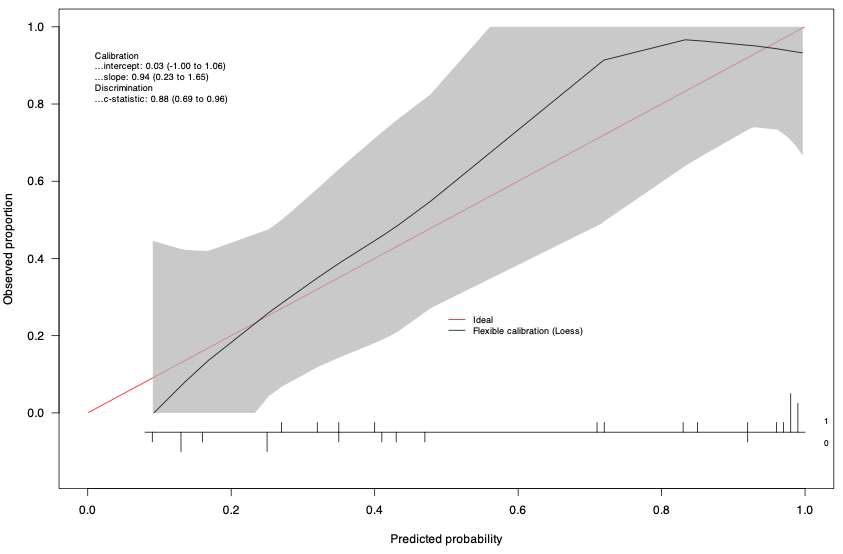

### **Appendix 11 – Decision curve analysis of the prediction model for typhoid incidence level, 2017-2024**

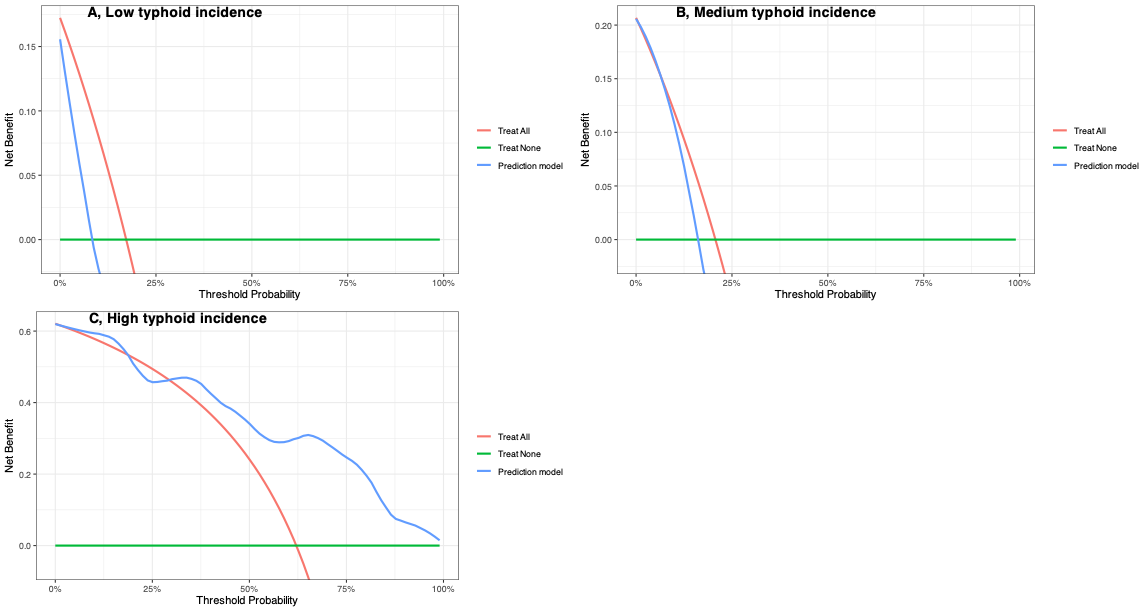

### **Appendix 12 - Simulated predicted probabilities for range of prevalence of *S.* Typhi among probable pathogens for typhoid incidence levels according to the final model**

| Prevalence of *S.* Typhi among probable pathogens, % | Predicted probability for low typhoid incidence site, % | Predicted probability for medium typhoid incidence site, % | Predicted probability for high typhoid incidence site, % | Suggested interpretation for typhoid incidence |
| --- | --- | --- | --- | --- |
| 1 | 62.3 | 27.9 | 9.8 | Low |
| 5 | 54.4 | 32.5 | 13.0 | Low |
| 10 | 44.3 | 37.3 | 18.4 | Low |
| 15 | 34.7 | 40.1 | 25.2 | Medium |
| 20 | 26.1 | 40.3 | 33.6 | Medium |
| 25 | 19.0 | 37.7 | 43.2 | High |
| 30 | 13.5 | 33.1 | 53.3 | High |
| 40 | 6.5 | 21.5 | 72.0 | High |
| 50 | 3.0 | 11.7 | 85.3 | High |

### **Appendix 13 - Metrics from sentinel bloodstream infection calculated for *S.* Paratyphi A and non-typhoidal *S.*, 2017-2024**

| **Study site:**  **Project, Country, Locality (reference)** | **Paratyphoid A incidence estimate per**  **100,000 person-years**  **(95% CI)** | | **Number of**  ***S.* Paratyphi A among**  **probable pathogens (%)** | | **Rank order of *S.***  **Paratyphi A** | ***S.* Paratyph*i* A / *E. coli* (ratio)** | | ***S.* Paratyphi A/**  **'stably endemic organisms'* (ratio)** | |
| --- | --- | --- | --- | --- | --- | --- | --- | --- | --- |
| Lao PDR,Vientiane ^1^ | 1 | (0-1) | 2/375 | (0.5) | 21 | 2/148 | (0.0) | 375/191 | (0.0) |
| Nepal,Lalitpur ^13^ | 33 | (12-72) | 4/57 | (7.0) | 2 | 4/2 | (2.0) | 57/3 | (1.3) |
| SEAP,Nepal,Kathmandu ^3^ | 81 | (56-118) | 42/236 | (17.8) | 2 | 42/11 | (3.8) | 236/11 | (3.8) |
| SEAP,Nepal,Kavrepalanchok ^3^ | 46 | (34-62) | 13/97 | (13.4) | 2 | 13/6 | (2.2) | 97/7 | (1.9) |
| SEAP,Pakistan,Karachi,AKUH ^3^ | 23 | (19-29) | 87/864 | (10.1) | 2 | 87/36 | (2.4) | 864/39 | (2.2) |
| SEAP,Pakistan,Karachi,KGH ^3^ | 1 | (1-1) | 2/644 | (0.3) | 3 | 2/2 | (1.0) | 644/3 | (0.7) |
| SEFI,India,Vellore ^4^ | 8 | (1-44) | 2/156 | (1.3) | 3 | 2/0 | - | 156/3 | (0.7) |

AKUH, Aga Khan University Hospital; CI, confidence interval; KGH, Kharadar General Hospital; SEAP, Surveillance for Enteric Fever in Asia Project; SEFI, Surveillance of Enteric Fever in India; *Stably endemic organisms were defined as *E. coli*, *S. pneumoniae,* or *S. aureus.*

| **Study site:**  **Project, Country, Locality (reference)** | **Non-typhoidal salmonella incidence estimate per**  **100,000 person-years**  **(95% CI)** | | **Number of**  **non-typhoidal salmonella among**  **probable pathogens (%)** | | **Rank order of non-typhoidal salmonella** | **Non-typhoidal salmonella*.* prevalence / *E. coli* prevalence (ratio)** | | **Non-typhoidal salmonella prevalence /**  **'stably endemic organisms' prevalence * (ratio)** | |
| --- | --- | --- | --- | --- | --- | --- | --- | --- | --- |
| TSAP,Burkina Faso,Nioko II ^6^ | 237 | (178-316) | 10/24 | (41.7) | 1 | 10/3 | (3.3) | 24/4 | (2.5) |
| TSAP,Burkina Faso,Polesgo ^6^ | 431 | (162-1147) | 4/27 | (14.8) | 3 | 4/4 | (1.0) | 27/4 | (1.0) |
| TSAP,Ethiopia,Butajira ^6^ | 0 |  | 0/12 | (0.0) | - | 0/2 | (0.0) | 12/6 | (0.0) |
| TSAP,Ghana,Asante Akim North ^6^ | 742 | (631-873) | 59/157 | (37.6) | 1 | 59/7 | (8.4) | 157/40 | (1.5) |
| TSAP,Guinea-Bissau,Bandim ^6^ | 37 | (24-57) | 8/20 | (40.0) | 1 | 8/0 |  | 20/6 | (1.3) |
| TSAP,Kenya,Kibera ^6^ | 32 | (14-70) | 6/107 | (5.6) | 4 | 6/4 | (1.5) | 107/47 | (0.1) |
| TSAP,Madagascar,Imerintsiatosika ^6^ | 9 | (2-50) | 1/10 | (10.0) | 3 | 1/1 | (1.0) | 10/2 | (0.5) |
| TSAP,Madagascar,Isotry ^6^ | 0 | (0-0) | 0/20 | (0.0) | - | 0/0 |  | 20/2 | (0.0) |
| TSAP,Senegal,Pikine ^6^ | 5 |  | 4/29 | (13.8) | 3 | 4/9 | (0.4) | 29/9 | (0.4) |
| TSAP,South Africa,Pietermaritzburg ^6^ | 0 |  | 0/36 | (0.0) | - | 0/5 | (0.0) | 36/22 | (0.0) |
| TSAP,Sudan,East Wad Medani ^6^ | 0 | (0-0) | 0/11 | (0.0) | - | 0/3 | (0.0) | 11/6 | (0.0) |
| TSAP,Tanzania,Moshi Rural District ^6^ | 7 | (2-23) | 1/11 | (9.1) | 4 | 1/4 | (0.3) | 11/6 | (0.2) |
| TSAP,Tanzania,Moshi Urban District ^6^ | 19 | (5-64) | 1/15 | (6.7) | 4 | 1/5 | (0.2) | 15/6 | (0.2) |

CI, confidence interval; TSAP, Typhoid Fever Surveillance in Africa Program. *Stably endemic organisms were defined as *E. coli*, *S. pneumoniae,* or *S. aureus.*

|  | **Meta-regression for log non-typhoidal *S.* incidence estimate** | **Univariate ordinal regression analysis for low, medium and high non-typhoidal *S.* incidence estimate levels^±^** |
| --- | --- | --- |
| **Metrics of sentinel bloodstream infection** | **Beta (95%CI)** | **OR (95%CI)** |
| Prevalence of non-typhoidal *S.* among probable pathogens (n=13) | 0.2 (0.08-0.33) | 1.12 (1.03-1.26) |
| Rank order of non-typhoidal *S.* among probable pathogens (n=9) | -0.98 (-2.7-0.73) | 0.43 (0.12-1.18) |
| Log non-typhoidal *S.* to *E. coli* ratio (n=11) | 1.16 (0.68-1.63) | 3.95 (1.32-30.6) |
| Log non-typhoidal *S.* to 'stably endemic organisms' ratio *(n=13) | 1.23 (0.76-1.69) | 3.40 (1.35-18.7) |

OR, odds ratio

± Low incidence (<10/100,000 person-years); medium incidence (10-100/100,000 person-years); high incidence (>100/100,000 person-years)

*Stably endemic organisms were defined as *E. coli*, *S. pneumoniae,* or *S. aureus.*
